# Exemplar Scoring Identifies Genetically Separable Phenotypes of Lithium Responsive Bipolar Disorder

**DOI:** 10.1101/2020.05.19.20098566

**Authors:** Abraham Nunes, William Stone, Raffaella Ardau, Anne Berghöfer, Alberto Bocchetta, Caterina Chillotti, Valeria Deiana, Franziska Degenhardt, Andreas J. Forstner, Julie S. Menzies, Eva Grof, Tomas Hajek, Mirko Manchia, Francis McMahon, Bruno Müller-Oerlinghausen, Markus M. Nöthen, Marco Pinna, Claudia Pisanu, Claire O’Donovan, Marcella D.C. Rietschel, Guy Rouleau, Thomas Schulze, Giovanni Severino, Claire M Slaney, Alessio Squassina, Aleksandra Suwalska, Gustavo Turecki, Petr Zvolsky, Pablo Cervantes, Maria del Zompo, Paul Grof, Janusz Rybakowski, Leonardo Tondo, Thomas Trappenberg, Martin Alda

**Affiliations:** Department of Psychiatry, Dalhousie University, Halifax, Nova Scotia, Canada; Faculty of Computer Science, Dalhousie University, Halifax, Nova Scotia, Canada; Unit of Clinical Pharmacology & San Giovanni di Dio Hospital, University Hospital of Cagliari, Cagliari, Italy; Charité University Medical Center, Campus Charité Mitte, Berlin, Germany; Department of Biomedical Sciences, Section of Neuroscience & Clinical Pharmacology, University of Cagliari, Cagliari, Italy; Institute of Human Genetics, University of Bonn, School of Medicine & University Hospital Bonn, Bonn, Germany; Centre for Human Genetics, University of Marburg, Marburg, Germany; Department of Biomedicine, University of Basel, Basel, Switzerland; Mood Disorders Center of Ottawa, Ottawa, Ontario, Canada; Department of Psychiatry, University of Toronto, Toronto, Ontario, Canada; Department of Medical Sciences and Public Health, Section of Psychiatry, University of Cagliari, Cagliari, Italy; Department of Pharmacology, Dalhousie University, Halifax, Nova Scotia, Canada; National Institute of Mental Health, Bethesda, MD, USA; Charité Universitätsmedizin-Berlin, Berlin, Germany; Centro Lucio Bini, Cagliari e Roma, Italy; Central Institute of Mental Health, Medical Faculty Mannheim, Heidelberg University, Germany; Montreal Neurological Institute, McGill University, Montreal, QC, Canada; Institute of Psychiatric Phenomics and Genomics, Munich, Germany; Department of Adult Psychiatry, Poznan University of Medical Sciences, Poznan, Poland; Department of Mental Health, Poznan University of Medical Sciences, Poznan, Poland; Department of Psychiatry, Charles University, Prague, Czech Republic; Department of Psychiatry, McGill University Health Centre, Montreal, Québec Canada; Department of Psychiatric Nursing, Poznan University of Medical Sciences, Poznan, Poland; Harvard Medical School and McLean Hospital, Boston, Massachusetts, USA

**Keywords:** Bipolar disorder, lithium response, machine learning, genomics, representational Rényi heterogeneity

## Abstract

Predicting lithium response (LiR) in bipolar disorder (BD) could expedite effective pharmacotherapy, but phenotypic heterogeneity of bipolar disorder has complicated the search for genomic markers. We thus sought to determine whether patients with “exemplary phenotypes”—those whose clinical features are reliably predictive of LiR and non-response (LiNR)—are more genetically separable than those with less exemplary phenotypes. We applied machine learning methods to clinical data collected from people with BD (n=1266 across 7 international centres; 34.7% responders) to compute an “exemplar score,” which identified a subset of subjects whose clinical phenotypes were most robustly predictive of LiR/LiNR. For subjects whose genotypes were available (n=321), we evaluated whether responders/non-responders with exemplary phenotypes could be more accurately classified based on genetic data than those with non-exemplary phenotypes. We showed that the best LiR exemplars had later illness onset, completely episodic clinical course, absence of rapid cycling and psychosis, and few psychiatric comorbidities. The best exemplars of LiR and LiNR were genetically separable with an area under the receiver operating characteristic curve of 0.88 (IQR [0.83, 0.98]), compared to 0.66 [0.61, 0.80] (p=0.0032) among the poor exemplars. Variants in the Alzheimer’s amyloid secretase pathway, along with G-protein coupled receptor, muscarinic acetylcholine, and histamine H1R signaling pathways were particularly informative predictors. In sum, the most reliably predictive clinical features of LiR and LiNR patients correspond to previously well-characterized phenotypic spectra whose genomic profiles are relatively distinct. Future work must enlarge the sample for genomic classification and include prediction of response to other mood stabilizers.

## 1. Introduction

Bipolar disorder (BD) is a severe lifelong illness characterized by recurrent manias, depressions, and a relatively high suicide risk [1,2]. Mood stabilizer initiation occurs approximately a decade after symptom onset, on average [3], and the trial-and-error process of pharmacological optimization for BD may lengthen this time. However, by predicting individuals’ mood-stabilizer response, this burden of untreated illness may be reduced.

Clinical data are currently the best lithium response predictors. Responders often have a completely episodic course with full inter-episode remissions, absence of rapid cycling, and family history of fully remitting BD (particularly the lithium responsive type) in a first degree relative [4,5]. This has motivated the search for strong genomic predictors of lithium response, but they remain elusive [6].

In large multi-site studies, lithium responder and non-responder groups may be too heterogeneous to classify robustly. However, it is possible that within this pooled group of heterogeneous subjects there exist more distinct “exemplars” of each phenotype, whose clinical profiles are consistent across sites, and who may be genomically more distinct. Our paper is thus motivated by two questions. First, can clinical presentation identify exemplars of lithium response and non-response? Second, are clinical exemplars of lithium response and non-response more genetically separable than their less exemplary counterparts?

Using the largest clinical database on lithium treatment in BD, we developed a method for rating the degree to which a subject is an exemplar of lithium response or non-response, respectively (an exemplar score). We hypothesized that the clinical differences between the best exemplars of lithium response and non-response would be reflective of factors previously associated with the “classical” bipolar phenotype. Finally, on a subset of subjects who were genotyped, we hypothesized that clinically exemplary responders and non-responders would be more accurately separable by application of a machine learning (ML) classifier to their genomic data (compared to their counterparts with low exemplar scores).

## 2. Methods

Clinical and genetic data were collected in the context of protocols approved by the Ethics Committee of the former Health Agency of Cagliari (now University Hospital Health Agency of Cagliari) for the Cagliari (University) and Centro Bini samples, and the research ethics boards of the Nova Scotia Health Authority, the McGill University Health Centre, the Royal Ottawa Hospital, and the University of Poznan.

Our analysis is split into two parts. In Part 1, we use a multi-centre database of clinical variables in order to derive a score that identifies subjects whose clinical phenotypes reliably predict lithium response/non-response. Part 2 uses a separate set of genomic data collected from a subset of subjects included in the clinical data from Part 1. In Part 2, we compare the ability to classify lithium response using those genetic data when they are stratified according to subjects’ *clinical* exemplar scores.

### 2.1. Part 1: Scoring and Characterization of Clinical Exemplars

#### 2.1.1. Data Collection

Clinical data collection procedures were described in Nunes et al. [7]. Data consisted of 180 variables recorded prior to instituting lithium maintenance therapy in 1266 people with BD across 7 sites internationally (Table 1). Response was evaluated after a minimum treatment duration of 1 year. Lithium response was defined as a score of ≥ 7 on the previously validated Alda scale [8].

**Table 1.** Description of constituent datasets. *Abbreviations:* number of patients (N), lithium responders (LR+), Cagliari (Centro Bini; CB), Cagliari (University; CU), International Group for the Study of Lithium (IGSLi), Maritimes (MAR), Ontario (ON), Poznan (POZ).

| Sample | N (LR+) | Description |
| --- | --- | --- |
| **CB** | 324 (21%) | Patients followed at the Mood Disorder Lucio Bini Center in Cagliari, Italy. Clinical data collection and response assessment was done by two psychiatrists. |
| **CU** | 206 (29%) | Patients in the long term treatment program at the Lithium Clinic of the Unit of the Clinical Pharmacology Center, University Hospital of Cagliari, Italy. Clinical data collection and response assessment was done by three psychiatrists and three clinical psychopharmacologists. |
| **IGSLi** | 70 (100%) | Patients recruited for a genetic study of lithium responsive bipolar disorder. [9] By design of that study, all patients were lithium responders. Clinical data collection and response assessment was done by three psychiatrists. |
| **MAR** | 343 (20%) | Patients followed by the Mood Disorders program at the Nova Scotia Health Authority and the Maritime Bipolar Registry. Clinical data collection and response assessment was done by two psychiatrists and two research nurses working in pairs. |
| **MTL** | 95 (16%) | Patients followed by the Mood Disorders Program at the McGill University Health Centre. Clinical data collection and response assessment was done by one psychiatrist. |
| **ON** | 117 (84%) | Patients from our earlier studies of lithium responsive bipolar disorder, [9,10] which, like the IGSLi sample, explains the greater proportion of responders. Clinical data collection and response assessment was done by three psychiatrists (including MA, who is now in the Maritimes). |
| **POZ** | 111 (53%) | Patients followed longitudinally by the Psychiatry Department at the University of Poznan, Poland. Clinical data collection and response assessment was done by two psychiatrists. |

#### 2.1.2. Exemplar Scoring Based on Clinical Predictors

Subjects who are most exemplary of their clinical phenotype should be classified accurately by models trained on data from any given site. Our overall exemplar scoring protocol thus involves (1) obtaining out-of-sample predictions of every subject’s class based on models trained on each individual site’s data, then (2) summarizing accuracy and level of agreement with which each subject was classified into a single value known as the exemplar score (Figure 1).

**Figure 1.**
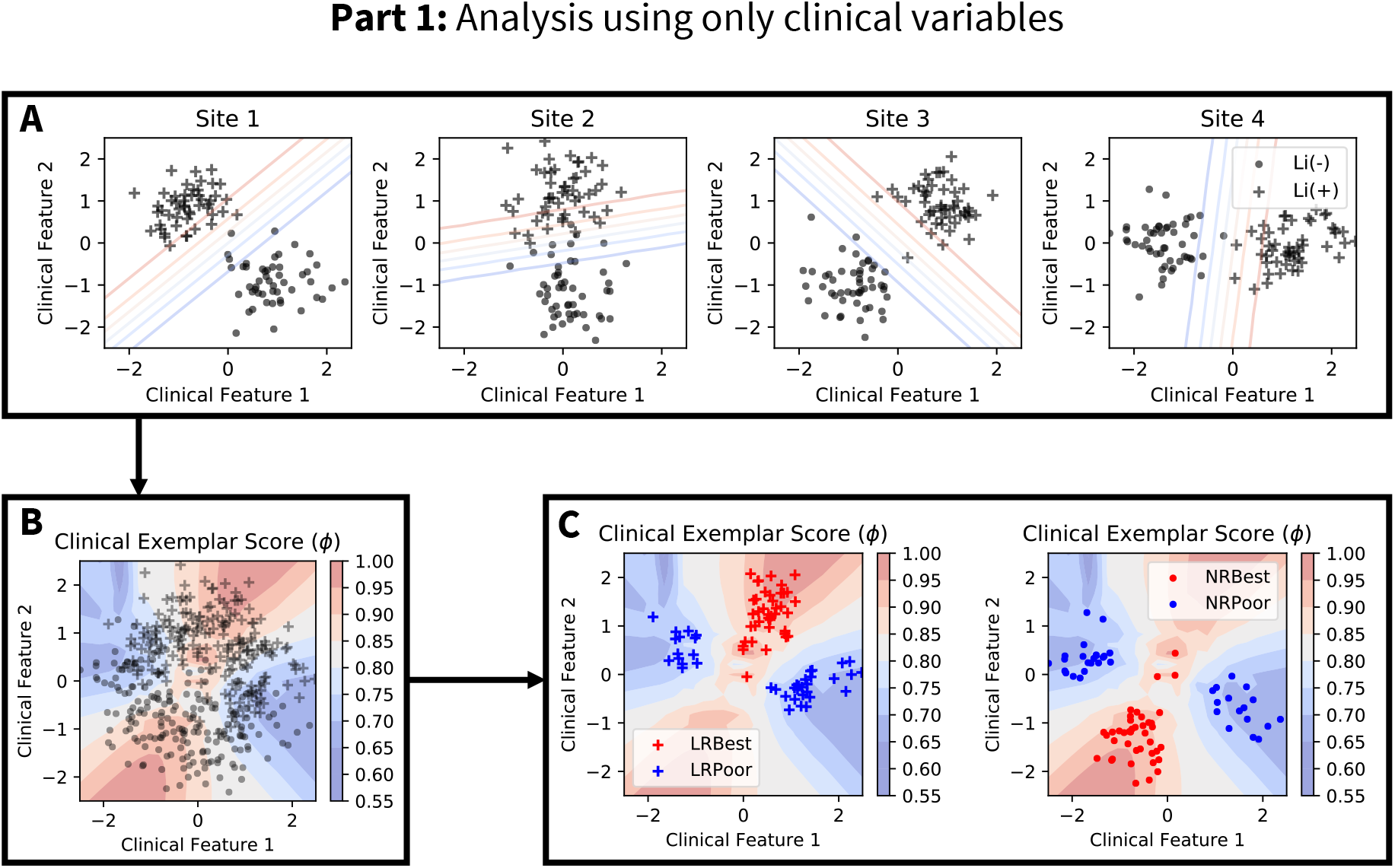
Hypothetical illustration of the clinical exemplar scoring analysis. Note that this part of the analysis is performed using the clinical feature dataset alone. **Panel A**: Demonstration of heterogeneity in the relationship between lithium responsiveness (depicted as “Li(+)” for responders and “Li(-)” for non-responders) and clinical features across four hypothetical sites. A classifier trained on data from each individual site may yield different discriminative functions. **Panel B**: Points demonstrate the aggregated dataset (“+” and are responders and non-responders, respectively). Contours demonstrate regions of clinical feature space in which site-level classifiers (from Panel A) agree with high accuracy on the predicted class. An exemplar score can be computed for each subject in the clinical dataset by (1) holding his data out of the training set, (2) predicting his lithium responsiveness using site-level classifiers trained on the remaining subjects, then (3) using the site-wise prediction results to compute the exemplar score. **Panel C**: Stratification of the clinical dataset according to lithium responsiveness and exemplar score quartile. The “LRBest” and “NRBest” exemplars are those responders and non-responders with exemplar scores above the 75th percentile, respectively. The “LRPoor” and “NRPoor” exemplars are those responders and non-responders with exemplar scores below the 25th percentile, respectively. This stratification can be used to evaluate the clinical features that differentiate good from poor exemplars of lithium response and non-response, respectively.

#### 2.1.3. The Clinical Exemplar Score

Let 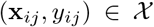 denote phenotypic data from subject *i ∊* {1, 2,…, *n_j_*}, where *x_ij_* is a vector of clinical features, *y_ij_* ∊ {0, 1} denotes whether the patient is a lithium responder, and *n_j_* is the number of patients in the sample from site *j* ∊ {1,2,…, *S*}. A pair (x, *y*) can thus be viewed as a set of coordinates on the (observable) phenotypic space 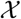. Data are sampled from *S* sites, each of which can be considered to sample a subdomain of the phenotypic space 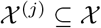. These site-wise subdomains are not necessarily disjoint. Indeed, if they were disjoint, the sites’ data would share nothing in common.

Now let 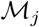 denote a classifier learned on training data from site *j*. Given a new set of clinical features, x′, the classifier predicts the probability that the corresponding patient is a lithium responder: that is, 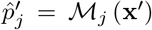. We denote the accuracy score of this prediction as

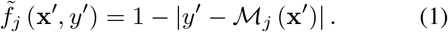

The representational Rényi heterogeneity measurement approach [11] consists of measuring heterogeneity on a latent or transformed space onto which observable data are mapped. To apply this in the present case, where we have defined our observable space, 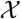, we must now devise an appropriate transformed space upon which the Rényi heterogeneity will be both meaningful and tractable. The heterogeneity deemed relevant in the present case arises in terms of differences in classification models across sites. Most starkly, we noted that the informative features for lithium response prediction varied between the best performing sites. In other words, depending on which site’s data are used for training, one might learn quite different (and perhaps even contradictory) relationships between clinical features and lithium responsiveness. In the limit where data from each site encodes completely different relationships between clinical features and lithium response, then each classifier 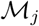 will behave distinctly (they will tend to disagree). In terms of numbers equivalent, we would say that in such a case there is an effective number of S distinct classifiers. Conversely, if the phenotypic domains of all sites overlap completely, then all classifiers 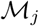 will tend to make similar predictions, which would correspond to an effective number of one classifier.

Let the accuracy of classifier 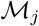 in predicting the relationship x → *y* be a measure of that model’s informativeness at point (x, *y*). We can thus define 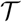 as a categorical space representing an index on “the most informative classifier.” We illustrate the mapping 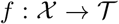 in Figure 2. A probability distribution over 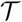 can be computed using a normalization of Equation 1:

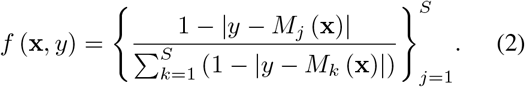

**Figure 2.**
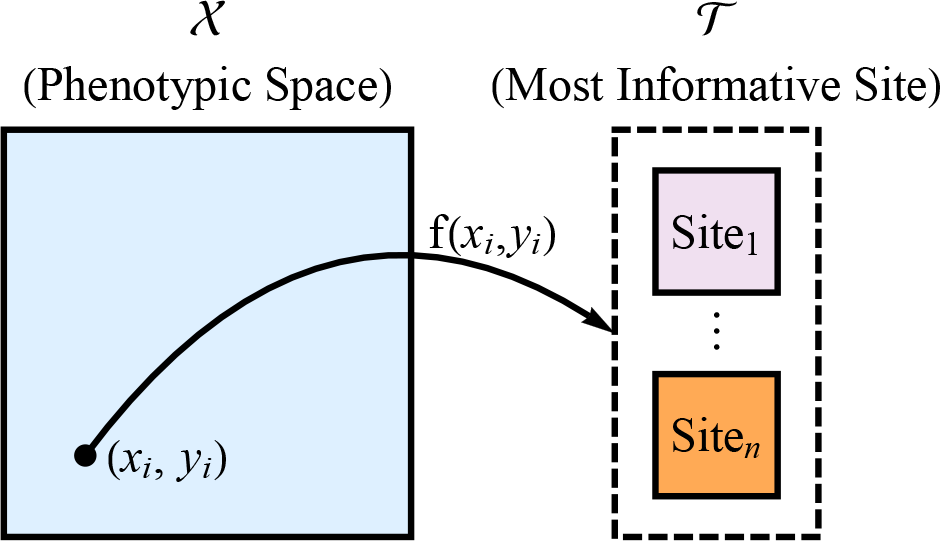
Representation of the mapping from phenotypic space 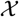 onto the representation of “most informative site-level model” (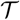). The transformation function is the normalized accuracy score for a classification model trained on each site’s data individually (Equation 2).

The quantity *f_j_* (x, *y*) can be taken to represent the probability that a classifier trained on data from site *j* is the most informative about the x → *y* mapping in that particular region of 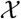. With this, we can compute the representational Rényi heterogeneity at (x, *y*) as follows:

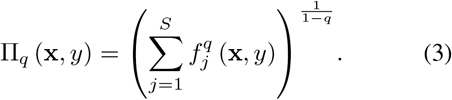

If the models 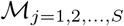 differ only in their training data (i.e. they have the same architecture, optimization routine, and hyperparameters) then the units of Equation 3 are “the effective number of informative sites.”

Recall that we defined a “clinical exemplar” as a subject whose phenotype (x, *y*) is reliably predicted accurately across all sites. In other words, regardless of the differences between sites’ data, all sites would agree in their predictions of the exemplars’ phenotypes. More formally, clinical exemplars must have high values of Π*_q_* (x, *y*) (all sites are similarly informative). However, to identify more specifically the exemplars of lithium response and non-response, we cannot solely rely on Π*_q_* (x, *y*), since that value may be high, despite sites’ prediction accuracies being low.

Let 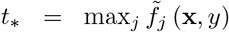 denote the maximal accuracy score obtained in classification at (x, *y*). We take this value to represent the degree to which a subject with that phenotype can be clearly associated with one class or another. An interesting case occurs where both *t*\* and Π*_q_* (x, *y*) are high, suggesting the point (x, *y*) is an exemplar of the regions of 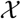 that are reliably well classified across sites. Conversely, if *t*_*_ ≈ 0.5 and Π*_q_* (x, *y*) is high, then that point is exemplary of a region of 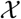 of which all sites are uncertain. When *t*\* is low and Π*_q_* (x, *y*) is high, then (x, *y*) is exemplary of a region of 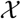 that reliably misleads all sites’ classifiers.

In the present study, we are concerned with identifying only those subjects with high values of both *t*\* and Π*_q_* (x, *y*), since they exemplify the most canonical “phenotypes” of lithium response and non-response, respectively. We accomplish this by combining t* and Π*_q_* (x, *y*) into a single index we call the *exemplar score*. The exemplar score at coordinate (x, *y*) of the phenotypic space is defined as

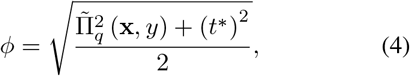

where 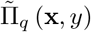 is a standardization of the Rényi heterogeneity to the [0,1] interval (the same scale as *t*\*):

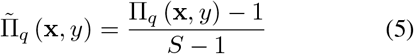

In the present study, we define the “best exemplars” as subjects whose exemplar scores (within their lithium response classes) were in the top 25%. Poor exemplars were those subjects whose phenotypes were in the lower quartile of exemplar scores within their response classes.

#### 2.1.4. The Predict Every Subject Out (PESO) Protocol

The predict every subject out (PESO) protocol is a method by which we can compute exemplar scores for each subject in the dataset while (A) ensuring that subject is not included in the training data and (B) having each model train on only that site’s data. All classifiers in our data were random forests, (RFC) [12] under the same specifications as in Nunes et al. [7] (100 estimators; SciKit Learn implementation; [13]). Similar to that study, missing data were marginalized by sampling from uninformative priors on respective variables’ domains [7]. A schematic of the protocol is shown in Figure 3.

For each site in the clinical predictors dataset, the PESO analysis protocol begins with a Leave-One-Out cross-validation run to obtain out-of-sample predictions for each of that site’s constituent subjects. We then train an RFC on that site’s data and predict lithium response in all other sites’ subjects. Each subject is thus mapped onto our categorical space 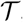T, upon which we can measure their exemplar scores.

**Figure 3.**
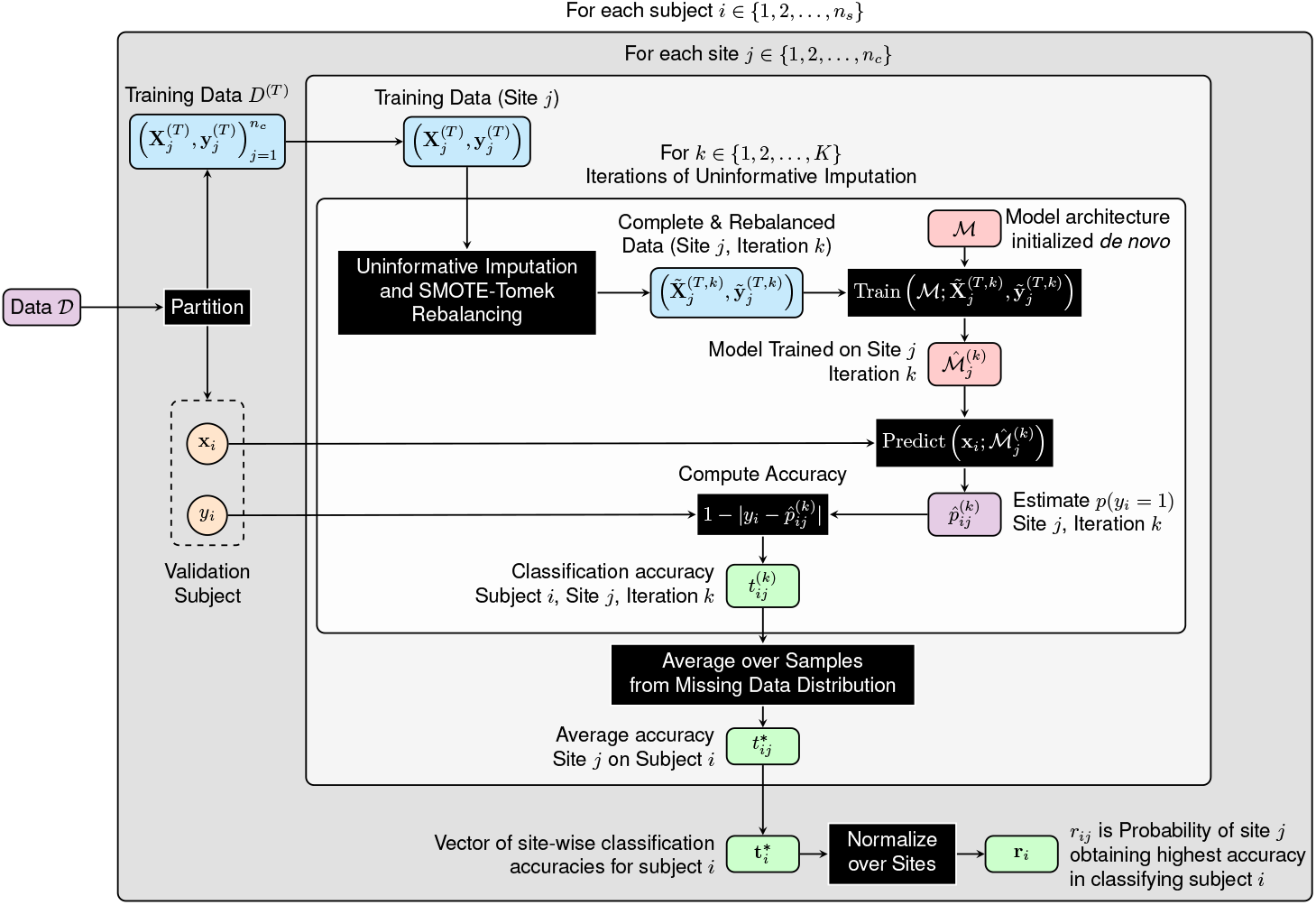
Illustration of the algorithm for the predict every subject out protocol.

#### 2.1.5. Comparison of Clinical Characteristics of the Best and Worst Exemplars

Univariate clinical feature differences were compared between the best exemplars of lithium response and non-response (“LRBest” and “NRBest,” respectively; the upper exemplar score quartile per class), and the corresponding poor exemplars (“LRPoor” and “NRPoor,” respectively; the lower exemplar score quartile per class). Continuous variables were compared using the two-sample permutation test of independence and categorical variables were compared using the randomization chi-square test (with 10,000 replications owing to multiple comparison corrections). The significance threshold was adjusted for 116 comparisons: *α_C_ =* 0.05/116 = 0.0004.

### 2.2. Part 2: Biological Validation hrough Genomic Classification

Figure 4 illustrates Part 2 of the present study, wherein we compare the genetic prediction of lithium response between subjects whose clinical profiles are exemplary and non-exemplary, respectively. After comparing genomic classification performance between the “Best” and “Poor” exemplar strata, respectively, we submit the genomic classifiers’ coefficients to gene enrichment analysis. This part of our study uses genomic data from subjects in the Consortium on Lithium Genetics GWAS cohort [6] who also had detailed clinical information collected for Part 1 of the present study.

**Figure 4.**
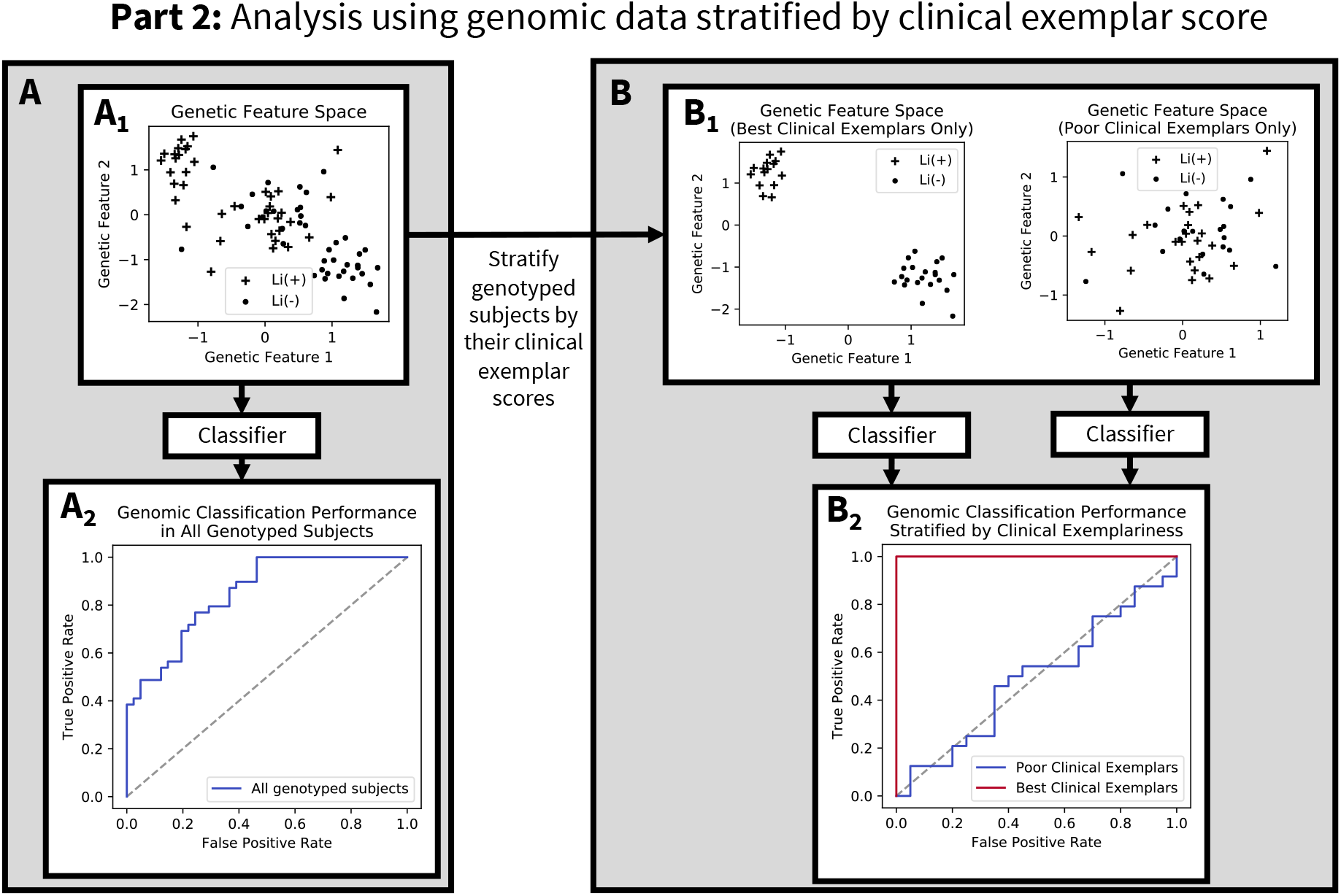
Hypothetical illustration of Part 2 of this study’s analysis, which evaluates the degree to which stratification of genomic data by corresponding subjects’ *clinical* exemplar scores can improve genomic classification performance. **Panel A**: Subjects’ genotypes lie on a genotypic feature space (shown in Panel A_1_ as a simplified 2 dimensional plane). Panel A_2_ shows a hypothetical ROC curve for these aggregated data. **Panel B:** Each genotyped subject has an exemplar score computed from Part 1 of the present study. Recall that the exemplar score merely identifies the degree to which a subject’s clinical profile (i.e. symptoms, family history, comorbidities, etc.) is reliably predictive of lithium responsiveness. Panel B_1_ shows that the aggregated genotyped sample can then be stratified into the “Best” clinical exemplars (subjects with top 25% of clinical exemplar scores within each of the responder and non-responder groups, respectively), and the “Poor” clinical exemplars (those with the lowest 25% of clinical exemplar scores in each responsiveness class). We then apply classifiers to the genomic data in each of these “Best Exemplar” and “Poor Exemplar” strata, respectively, and compare classification performance (Panel B_2_). The hypothetical receiver operating characteristic curve in Panel B_2_ reflects our hypothesis, that genetic classification of lithium response will be superior among the subgroup of Best clinical exemplars.

#### 2.2.1. Data Collection

Genomic data, obtained as part of the ConLiGen GWAS [6], were available for 321 of the subjects whose clinical data were analyzed in Part 1 of our study. In the Supplementary Materials, we show that there was no population stratification in this subsample, particularly in comparison to the broader ConLiGen sample. We restricted the data to only the 47,465 SNPs for which complete data were available across all ConLiGen sites. Preprocessing and quality control were done according to the Hou et al. [6] protocol.

#### 2.2.2. Genomic Classification Analysis

For genotyped subjects, we compared the performance of a classifier applied to (A) all 321 subject’s genomic data, (B) the worst exemplars’ genomic data, and (C) the best exemplars’ genomic data. We employed L2-penalized logistic regression (C=1 set *a priori*). Model criticism was performed under stratified-10-fold cross-validation.

Our primary outcome was the average cross-validated Matthews correlation coefficient (MCC), which is conservative under class imbalance. Classification performance differences were compared between conditions using the Kruskal-Wallis test. Where a statistically significant difference was observed (at *α* = 0.05), pairwise comparisons were done with the Mann-Whitney U tests (at threshold *α_C_* = 0.05/3 = 0.017). We secondarily report accuracy, area under the receiver operating characteristic curve (ROC-AUC), Cohen’s kappa, sensitivity, specificity, positive predictive value (PPV), and negative predictive value (NPV).

In the model trained on the best exemplars, we indexed variants whose logistic regression coefficients agreed in sign across all cross-validation folds, then applied a statistical enrichment test to the nearest associated genes using the PANTHER classification system v. 14.1 [14]. To evaluate the relationship between exemplar strata and enriched pathways, we repeated this analysis using logistic regression coefficients from the poor exemplar group. The threshold for statistical significance was set at *α_FDR_* = 0.05, where FDR indicates correction for false discovery rate. Further gene set analysis details are provided in Appendix B.

## 3. Results

### 3.1. Part 1: Scoring and Characterization of Clinical Exemplars

#### 3.1.1. Accuracy Distributions in the Predict Every Subject Out Analysis

A classifier trained on data from the Maritimes site achieved the highest mean overall accuracy (0.59, 95% confidence interval, CI, [0.58, 0.6]; Figure 5), which appeared largely driven by that site’s ability to accurately classify its own subjects (0.69 [0.66, 0.71]), and those from Montreal (0.71 [0.67, 0.75]). However, Figure 5 shows that site-level models’ accuracy distributions were highly variable in shape and modality, suggesting heterogeneous classification behaviour between sites.

**Figure 5.**
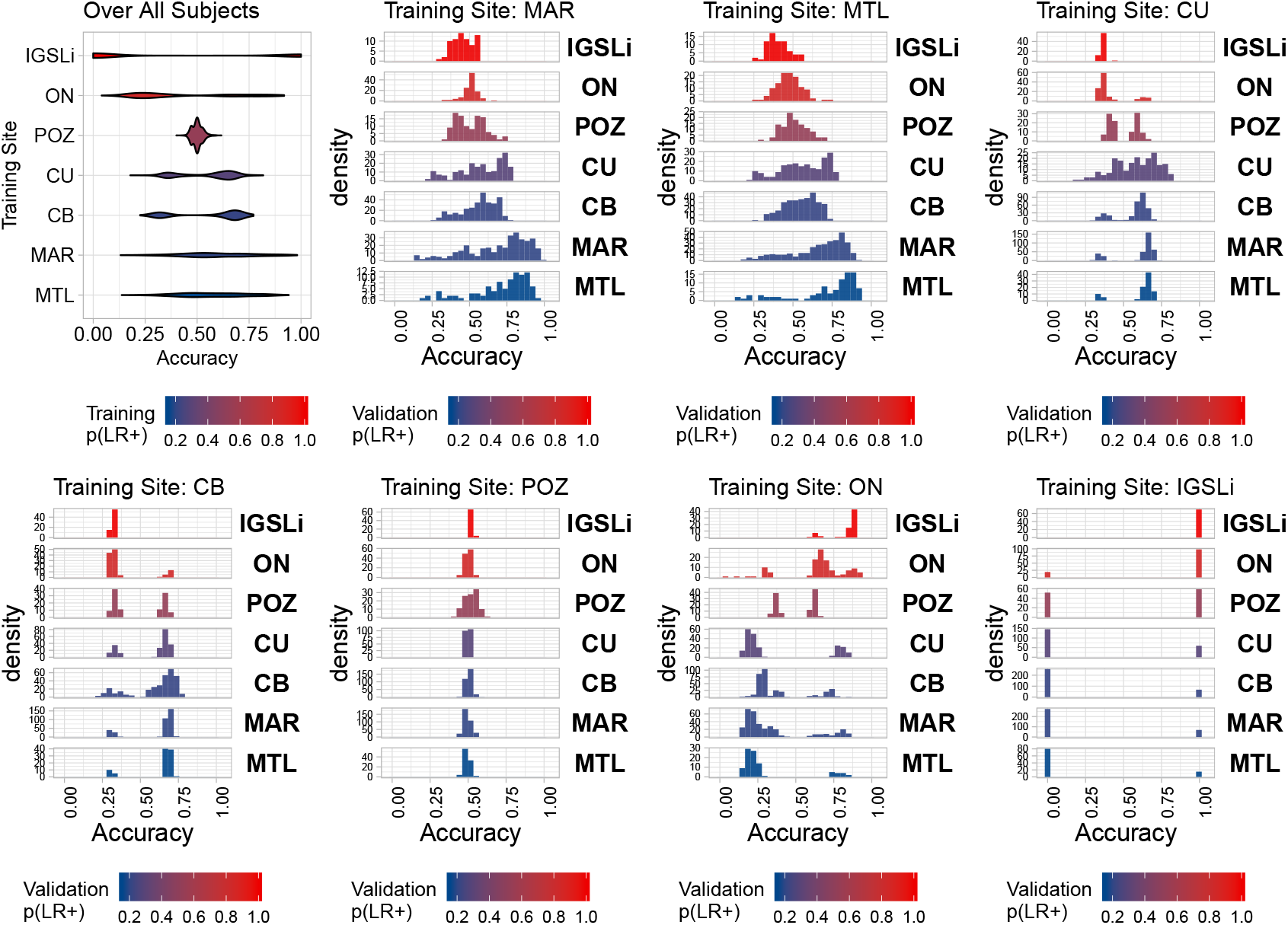
Accuracy distributions for models evaluated under the predict every subject out (PESO) regime. The violin plot at the upper leftmost corner shows the accuracy distributions for each site model evaluated over all subjects in the dataset, with the densities colored according to the proportion of lithium responders in the training site’s data. The remaining subplots show accuracy histograms for training site models (specified in the titles) stratified across out-of-sample sites. For the site-wise histograms, color indicates the responder/non-responder balance in the respective validation site. *Abbreviations*: Lithium responder (LR+), Cagliari (Centro Bini; CB), Cagliari (University; CU), International Group for the Study of Lithium (IGSLi), Maritimes (MAR), Ontario (ON), Poznan (POZ).

#### 3.1.2. Characteristics of the Best and Poor Exemplars

Within the clinical dataset of Part 1, there were 110 individuals in LRBest and LRPoor groups, and 207 individuals in the NRBest and NRPoor groups (Table 2). The LRBest group came predominantly from IGSLi (53.6%) and Ontario (21.8%), and most NRBest subjects were from Maritimes (72.5%) and Montreal (25.1%).

The LRBest group showed a later age of onset (median 28y, interquartile range, IQR [21, 36]) compared to NRBest (median 19, IQR [16, 24]; p<0.00001).

**Table 2.**
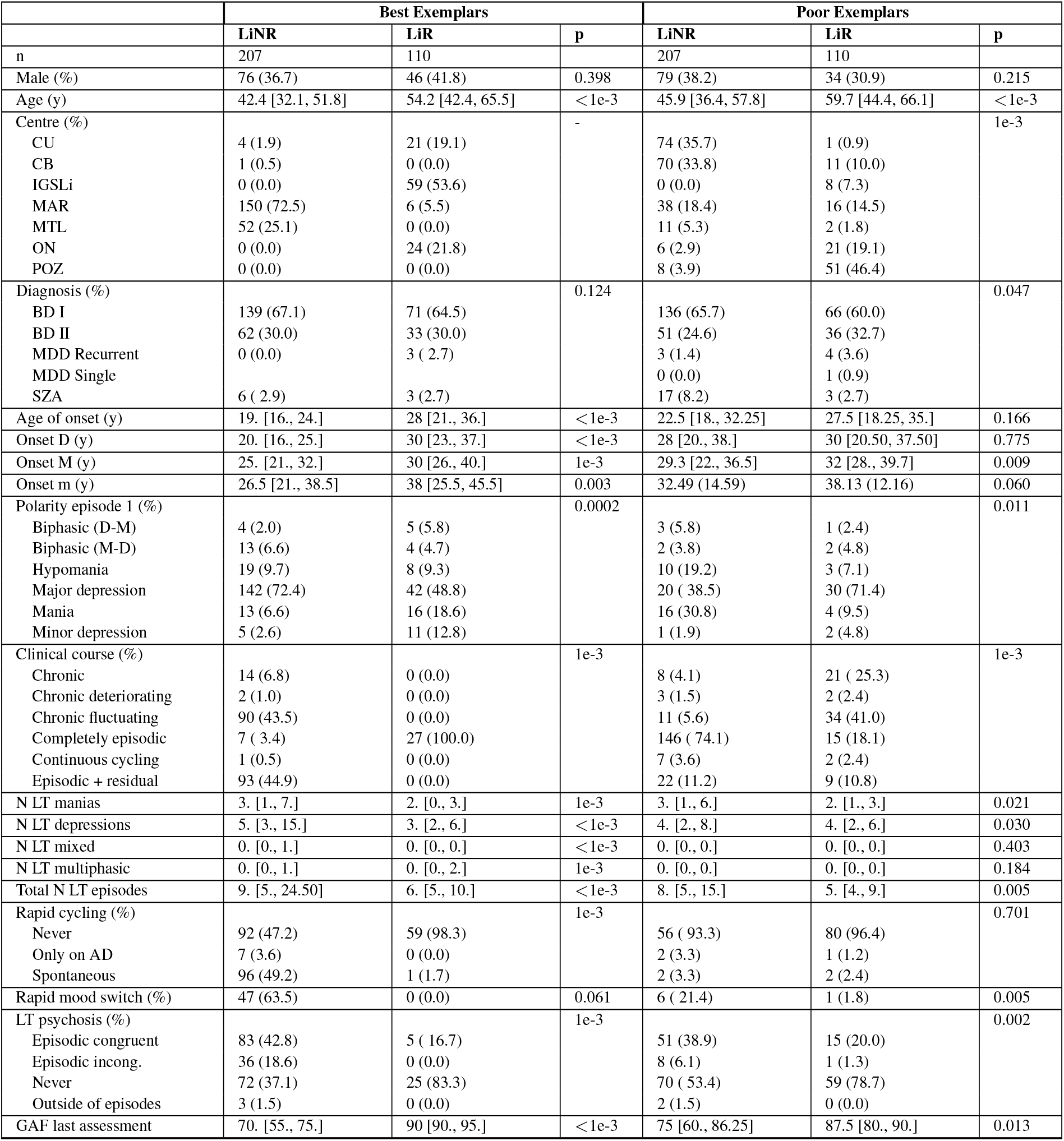
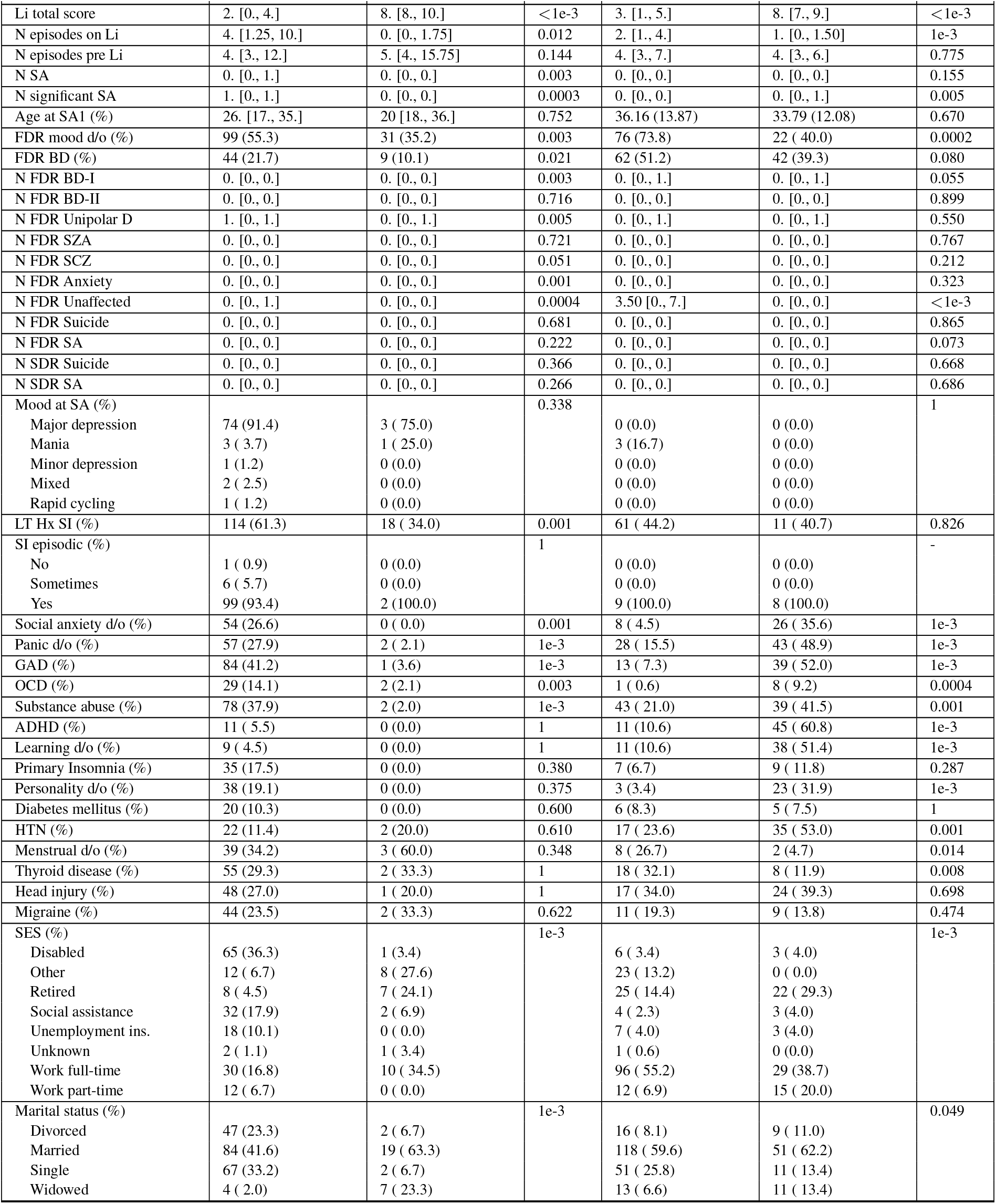
Clinical characteristics of exemplars, by lithium responsiveness. Characteristics of the best (upper 25% of exemplar scores) and poor (lower 25% of exemplar scores) exemplars of lithium response (LiR) and non-response (LiNR), respectively. Categorical data are presented as count (%), whereas normally distributed continuous variables are presented as mean (standard deviation), and non-normal continuous variables are presented as median [interquartile range]. Abbreviations: Calgiari (University; CU), Cagliari (Centro Bini; CB), International Group for the Study of Lithium (IGSLi), Maritimes (MAR), Montreal (MTL), Ontario (ON), Poznan (POZ), bipolar disorder (BD), major depressive disorder (MDD), antidepressants (AD), schizoaffective disorder (SZA), global assessment of functioning (GAF), lithium (Li), suicide attempts (SA), first degree relatives (FDR), second degree relatives (SDR), schizophrenia (SCZ), suicidal ideation (SI), history (Hx), generalized anxiety disorder (GAD), obsessive compulsive disorder (OCD), attention deficit hyperactivity disorder (ADHD), hypertension (HTN), socioeconomic status (SES).

|  | Best Exemplars |  |  | Poor Exemplars |  |  |
| --- | --- | --- | --- | --- | --- | --- |
|  | LiNR | LiR | p | LiNR | LiR | p |
| n | 207 | 110 |  | 207 | 110 |  |
| Male (%) | 76 (36.7) | 46 (41.8) | 0.398 | 79 (38.2) | 34 (30.9) | 0.215 |
| Age (y) | 42.4 [32.1, 51.8] | 54.2 [42.4, 65.5] | <1e-3 | 45.9 [36.4, 57.8] | 59.7 [44.4, 66.1] | <1e-3 |
| Centre (%) |  |  | - |  |  | 1e-3 |
| CU | 4 (1.9) | 21 (19.1) |  | 74 (35.7) | 1 (0.9) |  |
| CB | 1 (0.5) | 0 (0.0) |  | 70 (33.8) | 11 (10.0) |  |
| IGSLi | 0 (0.0) | 59 (53.6) |  | 0 (0.0) | 8 (7.3) |  |
| MAR | 150 (72.5) | 6 (5.5) |  | 38 (18.4) | 16 (14.5) |  |
| MTL | 52 (25.1) | 0 (0.0) |  | 11 (5.3) | 2 (1.8) |  |
| ON | 0 (0.0) | 24 (21.8) |  | 6 (2.9) | 21 (19.1) |  |
| POZ | 0 (0.0) | 0 (0.0) |  | 8 (3.9) | 51 (46.4) |  |
| Diagnosis (%) |  |  | 0.124 |  |  | 0.047 |
| BD I | 139 (67.1) | 71 (64.5) |  | 136 (65.7) | 66 (60.0) |  |
| BD II | 62 (30.0) | 33 (30.0) |  | 51 (24.6) | 36 (32.7) |  |
| MDD Recurrent | 0 (0.0) | 3 (2.7) |  | 3 (1.4) | 4 (3.6) |  |
| MDD Single |  |  |  | 0 (0.0) | 1 (0.9) |  |
| SZA | 6 (2.9) | 3 (2.7) |  | 17 (8.2) | 3 (2.7) |  |
| Age of onset (y) | 19. [16., 24.] | 28 [21., 36.] | <1e-3 | 22.5 [18., 32.25] | 27.5 [18.25, 35.] | 0.166 |
| Onset D (y) | 20. [16., 25.] | 30 [23., 37.] | <1e-3 | 28 [20., 38.] | 30 [20.50, 37.50] | 0.775 |
| Onset M (y) | 25. [21., 32.] | 30 [26., 40.] | 1e-3 | 29.3 [22., 36.5] | 32 [28., 39.7] | 0.009 |
| Onset m (y) | 26.5 [21., 38.5] | 38 [25.5, 45.5] | 0.003 | 32.49 (14.59) | 38.13 (12.16) | 0.060 |
| Polarity episode 1 (%) |  |  | 0.0002 |  |  | 0.011 |
| Biphasic (D-M) | 4 (2.0) | 5 (5.8) |  | 3 (5.8) | 1 (2.4) |  |
| Biphasic (M-D) | 13 (6.6) | 4 (4.7) |  | 2 (3.8) | 2 (4.8) |  |
| Hypomania | 19 (9.7) | 8 (9.3) |  | 10 (19.2) | 3 (7.1) |  |
| Major depression | 142 (72.4) | 42 (48.8) |  | 20 (38.5) | 30 (71.4) |  |
| Mania | 13 (6.6) | 16 (18.6) |  | 16 (30.8) | 4 (9.5) |  |
| Minor depression | 5 (2.6) | 11 (12.8) |  | 1 (1.9) | 2 (4.8) |  |
| Clinical course (%) |  |  | 1e-3 |  |  | 1e-3 |
| Chronic | 14 (6.8) | 0 (0.0) |  | 8 (4.1) | 21 (25.3) |  |
| Chronic deteriorating | 2 (1.0) | 0 (0.0) |  | 3 (1.5) | 2 (2.4) |  |
| Chronic fluctuating | 90 (43.5) | 0 (0.0) |  | 11 (5.6) | 34 (41.0) |  |
| Completely episodic | 7 (3.4) | 27 (100.0) |  | 146 (74.1) | 15 (18.1) |  |
| Continuous cycling | 1 (0.5) | 0 (0.0) |  | 7 (3.6) | 2 (2.4) |  |
| Episodic + residual | 93 (44.9) | 0 (0.0) |  | 22 (11.2) | 9 (10.8) |  |
| N LT manias | 3. [1., 7.] | 2. [0., 3.] | 1e-3 | 3. [1., 6.] | 2. [1., 3.] | 0.021 |
| N LT depressions | 5. [3., 15.] | 3. [2., 6.] | <1e-3 | 4. [2., 8.] | 4. [2., 6.] | 0.030 |
| N LT mixed | 0. [0., 1.] | 0. [0., 0.] | <1e-3 | 0. [0., 0.] | 0. [0., 0.] | 0.403 |
| N LT multiphasic | 0. [0., 1.] | 0. [0., 2.] | 1e-3 | 0. [0., 0.] | 0. [0., 0.] | 0.184 |
| Total N LT episodes | 9. [5., 24.50] | 6. [5., 10.] | <1e-3 | 8. [5., 15.] | 5. [4., 9.] | 0.005 |
| Rapid cycling (%) |  |  | 1e-3 |  |  | 0.701 |
| Never | 92 (47.2) | 59 (98.3) |  | 56 (93.3) | 80 (96.4) |  |
| Only on AD | 7 (3.6) | 0 (0.0) |  | 2 (3.3) | 1 (1.2) |  |
| Spontaneous | 96 (49.2) | 1 (1.7) |  | 2 (3.3) | 2 (2.4) |  |
| Rapid mood switch (%) | 47 (63.5) | 0 (0.0) | 0.061 | 6 (21.4) | 1 (1.8) | 0.005 |
| LT psychosis (%) |  |  | 1e-3 |  |  | 0.002 |
| Episodic congruent | 83 (42.8) | 5 (16.7) |  | 51 (38.9) | 15 (20.0) |  |
| Episodic incong. | 36 (18.6) | 0 (0.0) |  | 8 (6.1) | 1 (1.3) |  |
| Never | 72 (37.1) | 25 (83.3) |  | 70 (53.4) | 59 (78.7) |  |
| Outside of episodes | 3 (1.5) | 0 (0.0) |  | 2 (1.5) | 0 (0.0) |  |
| GAF last assessment | 70. [55., 75.] | 90 [90., 95.] | <1e-3 | 75 [60., 86.25] | 87.5 [80., 90.] | 0.013 |
|  | LiNR | LiR | p | LiNR | LiR | p |
| Li total score | 2. [0., 4.] | 8. [8., 10.] | < 1e-3 | 3. [1., 5.] | 8. [7., 9.] | < 1e-3 |
| N episodes on Li | 4. [1.25, 10.] | 0. [0., 1.75] | 0.012 | 2. [1., 4.] | 1. [0., 1.50] | 1e-3 |
| N episodes pre Li | 4. [3., 12.] | 5. [4., 15.75] | 0.144 | 4. [3., 7.] | 4. [3., 6.] | 0.775 |
| N SA | 0. [0., 1.] | 0. [0., 0.] | 0.003 | 0. [0., 0.] | 0. [0., 0.] | 0.155 |
| N significant SA | 1. [0., 1.] | 0. [0., 0.] | 0.0003 | 0. [0., 0.] | 0. [0., 1.] | 0.005 |
| Age at SA1 (%) | 26. [17., 35.] | 20 [18., 36.] | 0.752 | 36.16 (13.87) | 33.79 (12.08) | 0.670 |
| FDR mood d/o (%) | 99 (55.3) | 31 (35.2) | 0.003 | 76 (73.8) | 22 (40.0) | 0.0002 |
| FDR BD (%) | 44 (21.7) | 9 (10.1) | 0.021 | 62 (51.2) | 42 (39.3) | 0.080 |
| N FDR BD-I | 0. [0., 0.] | 0. [0., 0.] | 0.003 | 0. [0., 1.] | 0. [0., 1.] | 0.055 |
| N FDR BD-II | 0. [0., 0.] | 0. [0., 0.] | 0.716 | 0. [0., 0.] | 0. [0., 0.] | 0.899 |
| N FDR Unipolar D | 1. [0., 1.] | 0. [0., 1.] | 0.005 | 0. [0., 1.] | 0. [0., 1.] | 0.550 |
| N FDR SZA | 0. [0., 0.] | 0. [0., 0.] | 0.721 | 0. [0., 0.] | 0. [0., 0.] | 0.767 |
| N FDR SCZ | 0. [0., 0.] | 0. [0., 0.] | 0.051 | 0. [0., 0.] | 0. [0., 0.] | 0.212 |
| N FDR Anxiety | 0. [0., 0.] | 0. [0., 0.] | 0.001 | 0. [0., 0.] | 0. [0., 0.] | 0.323 |
| N FDR Unaffected | 0. [0., 1.] | 0. [0., 0.] | 0.0004 | 3.50 [0., 7.] | 0. [0., 0.] | < 1e-3 |
| N FDR Suicide | 0. [0., 0.] | 0. [0., 0.] | 0.681 | 0. [0., 0.] | 0. [0., 0.] | 0.865 |
| N FDR SA | 0. [0., 0.] | 0. [0., 0.] | 0.222 | 0. [0., 0.] | 0. [0., 0.] | 0.073 |
| N SDR Suicide | 0. [0., 0.] | 0. [0., 0.] | 0.366 | 0. [0., 0.] | 0. [0., 0.] | 0.668 |
| N SDR SA | 0. [0., 0.] | 0. [0., 0.] | 0.266 | 0. [0., 0.] | 0. [0., 0.] | 0.686 |
| Mood at SA (%) |  |  | 0.338 |  |  | 1 |
| Major depression | 74 (91.4) | 3 (75.0) |  | 0 (0.0) | 0 (0.0) |  |
| Mania | 3 (3.7) | 1 (25.0) |  | 3 (16.7) | 0 (0.0) |  |
| Minor depression | 1 (1.2) | 0 (0.0) |  | 0 (0.0) | 0 (0.0) |  |
| Mixed | 2 (2.5) | 0 (0.0) |  | 0 (0.0) | 0 (0.0) |  |
| Rapid cycling | 1 (1.2) | 0 (0.0) |  | 0 (0.0) | 0 (0.0) |  |
| LT Hx SI (%) | 114 (61.3) | 18 (34.0) | 0.001 | 61 (44.2) | 11 (40.7) | 0.826 |
| SI episodic (%) |  |  | 1 |  |  | - |
| No | 1 (0.9) | 0 (0.0) |  | 0 (0.0) | 0 (0.0) |  |
| Sometimes | 6 (5.7) | 0 (0.0) |  | 0 (0.0) | 0 (0.0) |  |
| Yes | 99 (93.4) | 2 (100.0) |  | 9 (100.0) | 8 (100.0) |  |
| Social anxiety d/o (%) | 54 (26.6) | 0 (0.0) | 0.001 | 8 (4.5) | 26 (35.6) | 1e-3 |
| Panic d/o (%) | 57 (27.9) | 2 (2.1) | 1e-3 | 28 (15.5) | 43 (48.9) | 1e-3 |
| GAD (%) | 84 (41.2) | 1 (3.6) | 1e-3 | 13 (7.3) | 39 (52.0) | 1e-3 |
| OCD (%) | 29 (14.1) | 2 (2.1) | 0.003 | 1 (0.6) | 8 (9.2) | 0.0004 |
| Substance abuse (%) | 78 (37.9) | 2 (2.0) | 1e-3 | 43 (21.0) | 39 (41.5) | 0.001 |
| ADHD (%) | 11 (5.5) | 0 (0.0) | 1 | 11 (10.6) | 45 (60.8) | 1e-3 |
| Learning d/o (%) | 9 (4.5) | 0 (0.0) | 1 | 11 (10.6) | 38 (51.4) | 1e-3 |
| Primary Insomnia (%) | 35 (17.5) | 0 (0.0) | 0.380 | 7 (6.7) | 9 (11.8) | 0.287 |
| Personality d/o (%) | 38 (19.1) | 0 (0.0) | 0.375 | 3 (3.4) | 23 (31.9) | 1e-3 |
| Diabetes mellitus (%) | 20 (10.3) | 0 (0.0) | 0.600 | 6 (8.3) | 5 (7.5) | 1 |
| HTN (%) | 22 (11.4) | 2 (20.0) | 0.610 | 17 (23.6) | 35 (53.0) | 0.001 |
| Menstrual d/o (%) | 39 (34.2) | 3 (60.0) | 0.348 | 8 (26.7) | 2 (4.7) | 0.014 |
| Thyroid disease (%) | 55 (29.3) | 2 (33.3) | 1 | 18 (32.1) | 8 (11.9) | 0.008 |
| Head injury (%) | 48 (27.0) | 1 (20.0) | 1 | 17 (34.0) | 24 (39.3) | 0.698 |
| Migraine (%) | 44 (23.5) | 2 (33.3) | 0.622 | 11 (19.3) | 9 (13.8) | 0.474 |
| SES (%) |  |  | 1e-3 |  |  | 1e-3 |
| Disabled | 65 (36.3) | 1 (3.4) |  | 6 (3.4) | 3 (4.0) |  |
| Other | 12 (6.7) | 8 (27.6) |  | 23 (13.2) | 0 (0.0) |  |
| Retired | 8 (4.5) | 7 (24.1) |  | 25 (14.4) | 22 (29.3) |  |
| Social assistance | 32 (17.9) | 2 (6.9) |  | 4 (2.3) | 3 (4.0) |  |
| Unemployment ins. | 18 (10.1) | 0 (0.0) |  | 7 (4.0) | 3 (4.0) |  |
| Unknown | 2 (1.1) | 1 (3.4) |  | 1 (0.6) | 0 (0.0) |  |
| Work full-time | 30 (16.8) | 10 (34.5) |  | 96 (55.2) | 29 (38.7) |  |
| Work part-time | 12 (6.7) | 0 (0.0) |  | 12 (6.9) | 15 (20.0) |  |
| Marital status (%) |  |  | 1e-3 |  |  | 0.049 |
| Divorced | 47 (23.3) | 2 (6.7) |  | 16 (8.1) | 9 (11.0) |  |
| Married | 84 (41.6) | 19 (63.3) |  | 118 (59.6) | 51 (62.2) |  |
| Single | 67 (33.2) | 2 (6.7) |  | 51 (25.8) | 11 (13.4) |  |
| Widowed | 4 (2.0) | 7 (23.3) |  | 13 (6.6) | 11 (13.4) |  |

The LRBest subjects for whom clinical course information was available all showed a completely episodic course, whereas NRBest courses were mainly chronic fluctuating (43.5%) and episodic with residual symptoms (44.9%). These differences were statistically significant at the omnibus level (p=0.0001). Interestingly, differences in clinical course between LRPoor and NRPoor were opposite in direction to those observed among best exemplars. NRPoor subjects had predominantly completely episodic clinical courses (74.1%), whereas LRPoor subjects exhibited predominantly chronic fluctuating (41%) and chronic (25.3%) courses, with only 18.1% being completely episodic (omnibus p=0.0001).

The complete absence of rapid cycling was reported in 98.3% of LRBest, and in only 47.2% of NRBest (p=0.0001). The remaining majority of the NRBest subjects (49.2%) reported having experienced spontaneous rapid cycling. The occurrence of rapid cycling was no different between LRPoor and NRPoor groups.

The occurrence of lifetime psychosis differed between LRBest and NRBest, with a total of 42.8% of the non-responders reporting episodic and mood congruent psychosis (compared to only 16.7% of responders; p=0.0001). Non-responders also reported incongruent episodic psychosis in 18.6% of cases, with only 37.1% of non-responders reporting an absence of psychosis altogether. In contrast, 83.3% of the best exemplars of lithium response reported a complete absence of lifetime psychosis.

The LRBest group had a lower rate of panic disorder (2.1% vs. 27.9%; p=0.0001), generalized anxiety disorder (3.6% vs 41.2%; p=0.00025), and substance abuse (2% vs. 37.9%; p=0.0001) than NRBest. There was also a general trend toward lower rates of psychiatric comorbidity in LRBest compared to the NRBest group. Social anxiety disorder was present in 0% of lithium responders but 27.9% of non-responders (p=0.0007). Responders also had relatively lower rates of obsessive-compulsive disorder (2.1%) compared to non-responders (14.1%; p=0.0025). These findings were largely reversed when looking at the poor exemplars. LRPoor subjects had higher rates of social anxiety disorder (35.6% vs 4.5%; p=0.0001), panic disorder (48.9% vs 15.5%; p=0.0001), generalized anxiety disorder (52% vs 7.3%; p=0.0001), substance abuse (41.5% vs 21.0%; p=0.0005), attention deficit hyperactivity disorder (60.8% vs. 10.6%; p=0.0001), learning disability (51.4% vs 10.6%; p=0.0001), and personality disorder (31.9% vs 3.4%; p=0.0001) compared to the NRPoor subjects.

### 3.2. Part 2: Biological Validation through Genomic Classification

Recall that the genomic data for this element of the analysis are derived from a single site in the ConLiGen data. In Appendix C, we demonstrate relative lack of genomic population stratification in this subset, with a comparison to the broader ConLiGen sample.

#### 3.2.1. Genomic Classification among the Best and Poor Exemplars

Genotyped subjects overlapped with clinical data from the Maritimes (n=129; 40%), Montreal (n=74; 23%), Ontario (n=62; 19%), and IGSLi (n=56; 17%), although in the ConLiGen GWAS [6], they were all classified as from the Maritimes (Dalhousie University). Most clinical differences reflect those reported in Section 3.1.2 and thus are reported in Table D.1.

Genomic classification results are presented in Figure 6 and in tabular fashion in Table D.2. The median MCC for classification of the Best exemplars was 0.58 (IQR [0.41, 0.77]), which was greater than classification analyses with either the poor exemplars (0.29 [0.06, 0.5]; p=0.0043), or the entire dataset (0.32 [0.2, 0.44]; p=0.002). The ROC-AUC for classification of lithium response in the Best exemplars was 0.88 [0.83, 0.98], which was greater than that of the model trained only on poor exemplars (0.66 [0.61, 0.80]; p=0.0032) or the whole dataset (0.7 [0.62, 0.75]; p=0.001).

**Figure 6.**
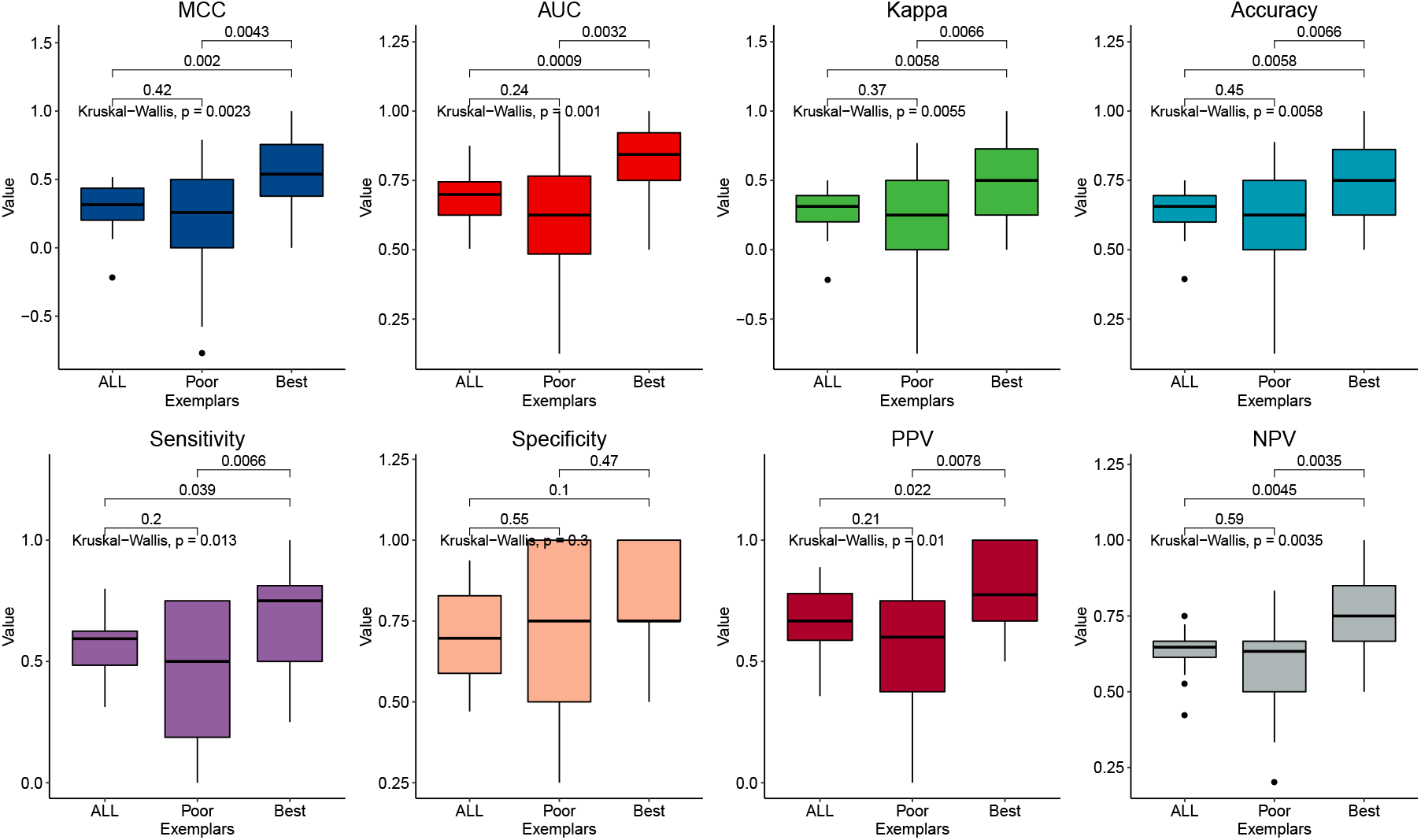
Genomic classification results. Results of classifying lithium response based on the genomic data of all subjects (“ALL”; n=321), the poor exemplars (<25th percentile of exemplar score; n=81), and the best exemplars (>75th percentile of exemplar score; n=79). Boxes are defined by the interquartile range (IQR), with the median shown as the black centered line. Whiskers are 1.5 times the IQR. Each panel shows the results for a different classification performance metric. *Abbreviations:* Matthews’ correlation coefficient (MCC), area under the receiver operating characteristic curve (AUC), Cohen’s kappa (Kappa), positive predictive value (PPV), negative predictive value (NPV).

Figure 7 shows pathway analysis results for the best exemplars. Enriched pathways involved (A) muscarinic acetylcholine receptor types 1 and 3 signaling (mAChR1/3; 27 genes, false discovery rate FDR=0.017), (B) Alzheimer disease-amyloid secretase (30 genes, FDR=0.034), (C) heterotrimeric G-protein coupled receptor Gq/Go α signaling (GPCRq/o-α; 53 genes, FDR=0.04), and (D) histamine H1R mediated signaling (H1R; 27 genes, FDR=0.039). Complete gene set analysis results are shown in Table D.3. Enrichment studies in the gene ontology “cellular component” and “biological function” categories are shown in Tables D.4 and D.5.

**Figure 7.**
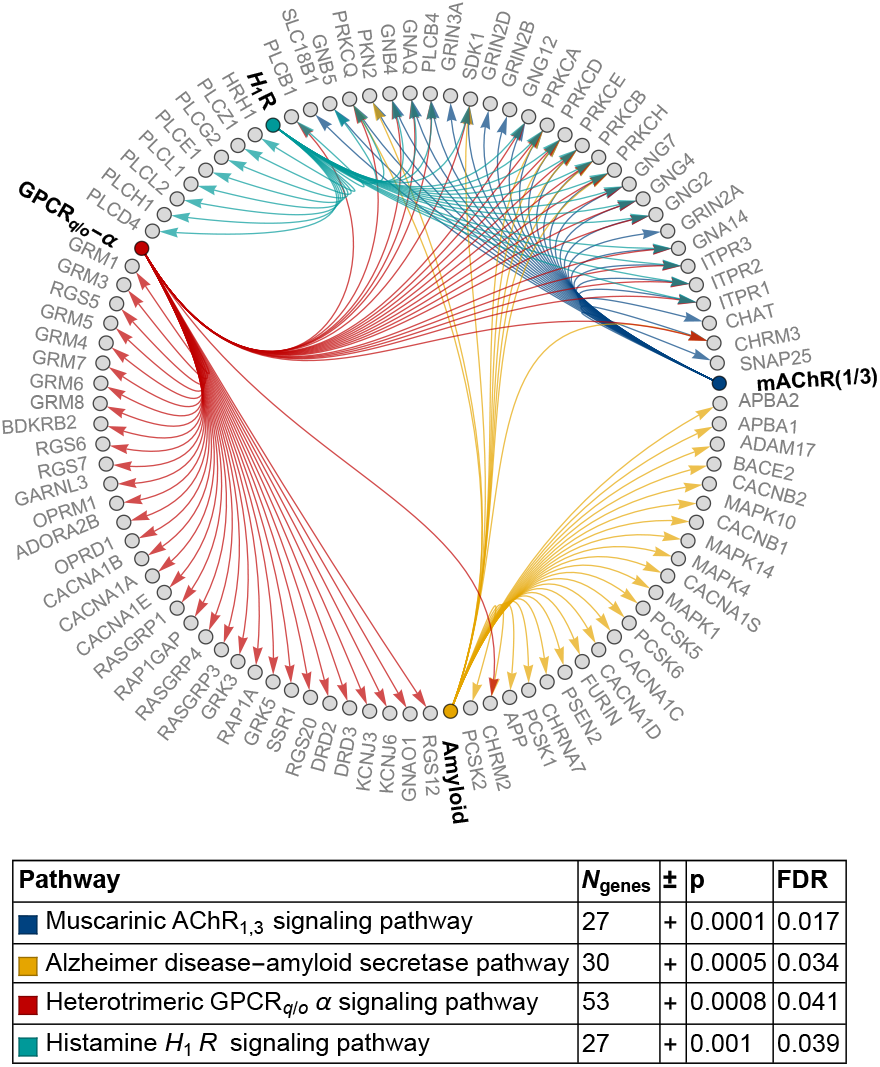
Gene enrichment in the best exemplars. Results of the statistical enrichment test using the logistic regression coefficients from the classifier trained on the best exemplar. Individual genes are shown in gray, with pathway nodes (and edges) colored according to the pathway identity. Pathway names are shown in bold along the perimeter of the graph. *Abbreviations:* acetylcholine receptor (AChR), G-protein coupled receptor (GPCR), histamine H1 receptor (H1R), false discovery rate (FDR).

## 4. Discussion

Individuals who are most phenotypically representative of lithium response and non-response may be more genetically distinct than their less exemplary counterparts, particularly in genes related to GPCRq/o-α, mAChR1/3 or H1R signaling, and the Alzheimer’s amyloid-secretase pathway. Exemplars also showed distinct clinical profiles that are consistent with past phenotypic research on lithium responders. Since clinical exemplars are more genetically separable, our study confers a measure of biological validity upon the practice of detailed clinical evaluation, whose predictive utility we have previously demonstrated [7].

One of our most important findings was characterization of the LRBest group as individuals with (A) a predominantly completely episodic clinical course, (B) low levels of psychiatric comorbidity, (C) later age of onset, (D) a general absence of rapid cycling, and (E) either absence of psychosis or limitation to mood congruent intra-episodic form. The first two findings are likely the strongest since we observe the opposite pattern among the LRPoor and NRPoor groups. Notwithstanding, all of these elements support past evidence on the clinical phenotype of lithium responsive bipolar disorder. For instance, Passmore et al. [15] found that lithium responders generally had a more episodic course of illness, whereas lamotrigine responders were more likely to have experienced rapid cycling, a higher rate of psychiatric comorbidity, and an earlier age of onset. A later age of onset in lithium responders has been demonstrated in meta-analysis [16,17]. Absence of rapid cycling has also been associated with good lithium response by Backlund et al [18] and Tondo et al. [19]. Finally, Kleindienst & Greil [20] found that carbamazepine responders were more likely to have had mood incongruent psychosis than lithium responders, while the updated meta-analysis by Hui et al. found an association between absence of psychotic symptoms and lithium responsiveness [17]. Aside from not including family history related variables (potentially an artifact of related variable definitions), the clinical picture of the exemplary lithium responder that emerges from our study largely aligns with that noted by several authors, such as Grof [4], Gershon & Malhi [21], and Alda [22].

Recently, Kendler [23] reminded us that the utility of biological tests, such as the electrocardiograms and troponin assays used to detect myocardial infarction, is generally contingent upon the clinician’s identification of candidate patients whose presentations are clinically consistent with the illness being targeted. The present study, which shows that refinement of a clinical sample into those whose phenotypes are clinically most exemplary of the target syndrome, provides strong data-driven support for Kendler’s statement. Further still, we have noted that the clinical picture of the exemplary lithium responders (and non-responders) has been hypothesized for some time, and our study now provides biological support for the predictive validity of these phenotypic hypotheses. Specifically, we were able to genomically classify the best clinical exemplars of lithium response and non-response with a ROC-AUC of 0.88 (IQR [0.83,0.98]), whereas poor exemplars of these classes could only be discriminated with a ROC-AUC of 0.66 (IQR [0.61,0.80]; p=0.0032). If there was no biologically mediated information in the exemplary phenotype of lithium response (and non-response), then this difference would not have been observed.

Variants most informative in discrimination of the best exemplars showed enrichment of genes involved in the heterotrimeric GPCRq/o-*α*, mAChR1/3 or H1R signaling, and the Alzheimer’s amyloid-secretase pathway. Lithium response and BD have long been associated with GPCR signaling [24]. In particular, lithium may affect signaling in both the Go-alpha pathway (at least via adenylate cyclase) and the Gq-alpha pathway (via effects on 1,4,5-triphosphate and protein kinase C, PKC) [25–29]. Interestingly, our results imply that differences in GPCR signaling may be segregated according to medication responsiveness. Enrichment in the Alzheimer’s amyloid-secretase pathway is interesting given the growing interest in the effects of lithium on Alzheimer’s pathology. Alterations in cholinergic and histaminergic systems have figured less prominently in the biological literature on BD and lithium response. However, note that Figure 7 shows that many genes enriched in the cholinergic and histaminergic systems were also enriched in the GPCR and Alzheimer’s amyloid pathways (which comparatively have more individual genetic associations). It is possible that alterations in the cholinergic and histaminergic systems may be subcomponents of the broader differences in the GPCR and Alzheimer’s amyloid systems. In future work, it would be of interest to characterize a more fine-grained “gradient” of genetic differences across the spectrum of exemplar scores, and to further evaluate the significance of cholinergic and histaminergic system enrichment in our study.

One limitation of our study includes the relatively low sample size for the genomic analysis. Future work could endeavor to obtain further genotypic information for individuals in our clinical database, or detailed clinical information for individuals in our genomic database. As features, our study also only used those SNPs that overlapped across genotyping platforms in the ConLiGen dataset. Unfortunately, however, the number of fully imputed variants was on the order of millions, which would be analytically intractable in the present context.

Filtering-based feature selection approaches in our present study would be (A) too computationally expensive across these millions of variants and (B) require much larger sample sizes since they must be repeated within each training partition. We also had no dominant a priori biological rationale for limiting the data to a restricted subset, since, as our results later confirmed, these biological systems may differ between exemplar strata. Ultimately, we chose the set of completely genotyped SNPs that overlapped across ConLiGen sites in order to facilitate the potential conceptual generalizability of our pathway analysis results, in particular. That is, since the pathways detected were based on variants that are broadly genotyped, these results could potentially be extended to other ConLiGen sites, should the corresponding clinical variables become available.

Our study is also limited by its focus on lithium response, at the exclusion of other mood stabilizers. It is therefore possible our lithium responders are simply those with a more generally responsive form of BD. The only way to prove specificity would be to obtain data showing a single subject’s non-response to other mood stabilizers and response to lithium. That being said, there is evidence that excellent response to lithium may be exclusive to that medication [29]. After further validity checks on larger samples of genomic data in lithium responders and non-responders, it will be of great interest to examine exemplar-based genomic classification of mood stabilizer response more broadly. Such work could potentially advance the development of joint clinical-biological prediction models for mood stabilizer response.

## Data Availability

Data requests may be provided to the authors.

## Funding

Genome Canada (MA, AN), Dalhousie Department of Psychiatry Research Fund (MA, AN), Dalhousie Medical Research Foundation and the Lindsay Family (MA, AN), Canadian Institutes of Health Research #418254 (MA, AN), Nova Scotia Health Research Foundation Scotia Scholars Graduate Scholarship (AN), Killam Postgraduate Scholarship (AN), EMBED-BMBF-01EW1904 (MDCR).

## Appendix B Gene Set Analysis

At each fold of cross-validation (under all settings of q), the logistic regression coefficients were saved. The SNPs whose logistic regression coefficients were of the same sign (i.e. positive or negative) across all folds were ranked in terms of their absolute median coefficient values and linked to gene identifiers using the NCBI gene database. Each gene was assigned the maximal absolute value of the logistic regression coefficients for all SNPs tagged by that gene; the remainder (duplicates) were deleted, such that each included gene had only one numerical value associated with it. We then applied the statistical enrichment test in the PANTHER classification system v. 14.1 [14]. We repeated the statistical enrichment test for the following annotation sets: PANTHER pathways, GO molecular function (complete), GO biological processes (complete), GO cellular components (complete). To further evaluate the degree to which the enrichment analyses speak specifically to findings among the best exemplars, we repeated the same procedures outlined here using the logistic regression coefficients for the poor exemplars.

## Appendix C Population Stratification

To evaluate for the presence of population stratification in our genomic sample, we plot the first several principal components of the subjects’ genotypes in Figure C.1. For comparison, Figure C.2 demonstrates the first several principal components from 14 sites of the full Consortium on Lithium Genetics (ConLiGen) genomic sample.

**Figure C.1.**
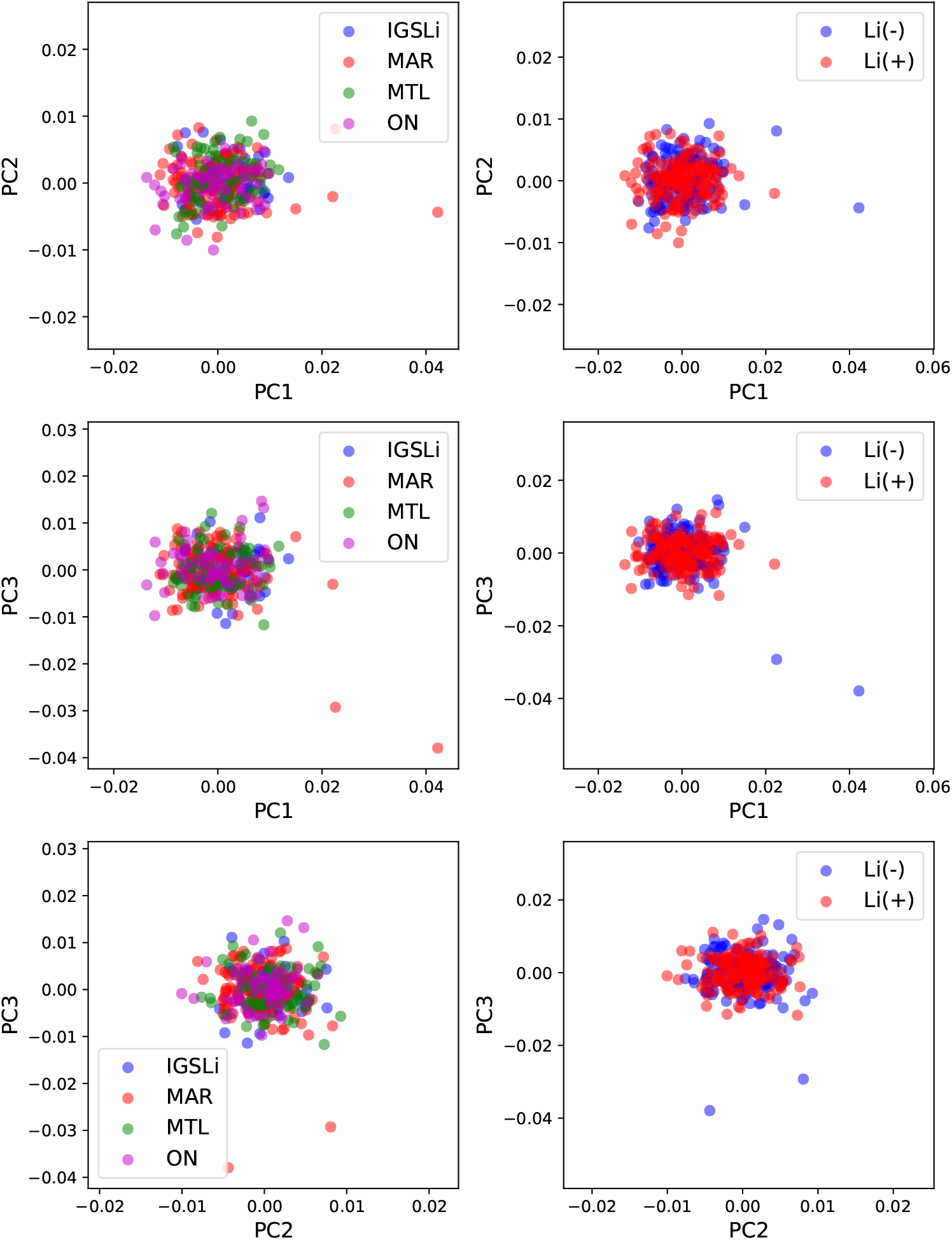
Principal components analysis of the genomic dataset from Halifax (as coded in the ConLiGen studies [6]). The left column is coloured by the site of origin, whereas the right column of plots is coloured by lithium responsiveness. *Abbreviations:* International Group for the Study of Lithium (IGSLi), Maritimes (MAR), Montreal (MTL), Ontario (ON; also known as Ottawa/Hamilton).

**Figure C.2.**
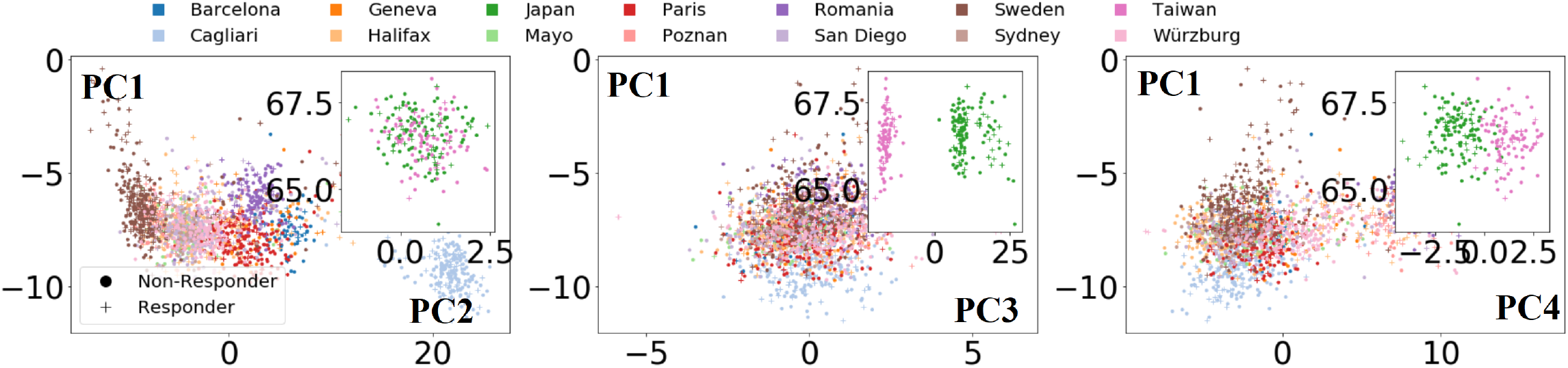
Principal components analysis of the genomic dataset from the Consortium on Lithium Genetics sample [6] (Figure used with permission from Stone et al., submitted manuscript)

## Appendix D Supplementary Tables

Clinical demographic comparisons between the best exemplars, poor exemplars, and the aggregated sample of genotyped patients is presented in Table D.1, with stratification by lithium response. The results of gene enrichment analysis are presented in Table D.3, with specific genes enriched in the best exemplar group (related to glutamate receptors and signalling processes) shown in Tables D.4 and D.5.

**Table D.1.**
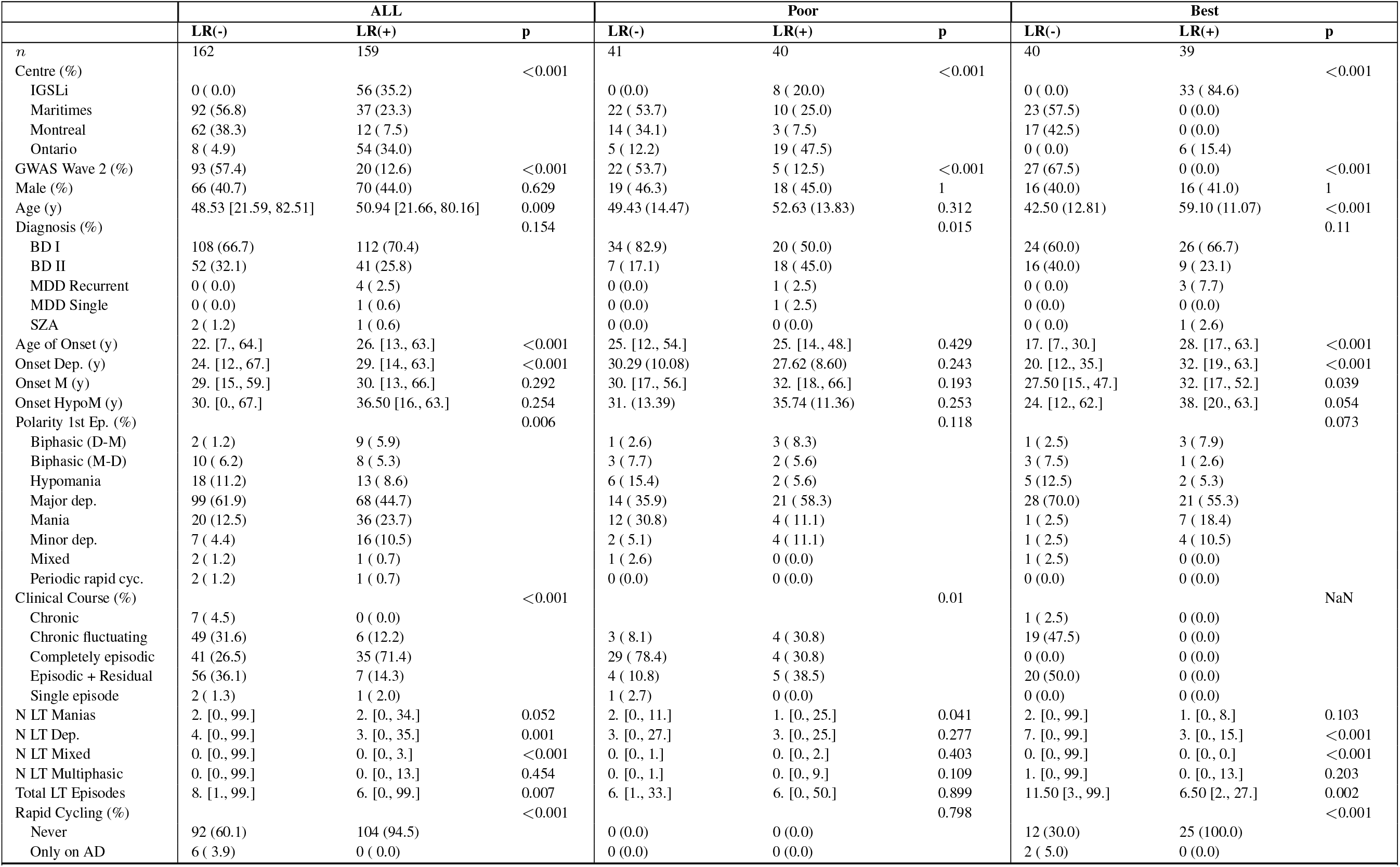
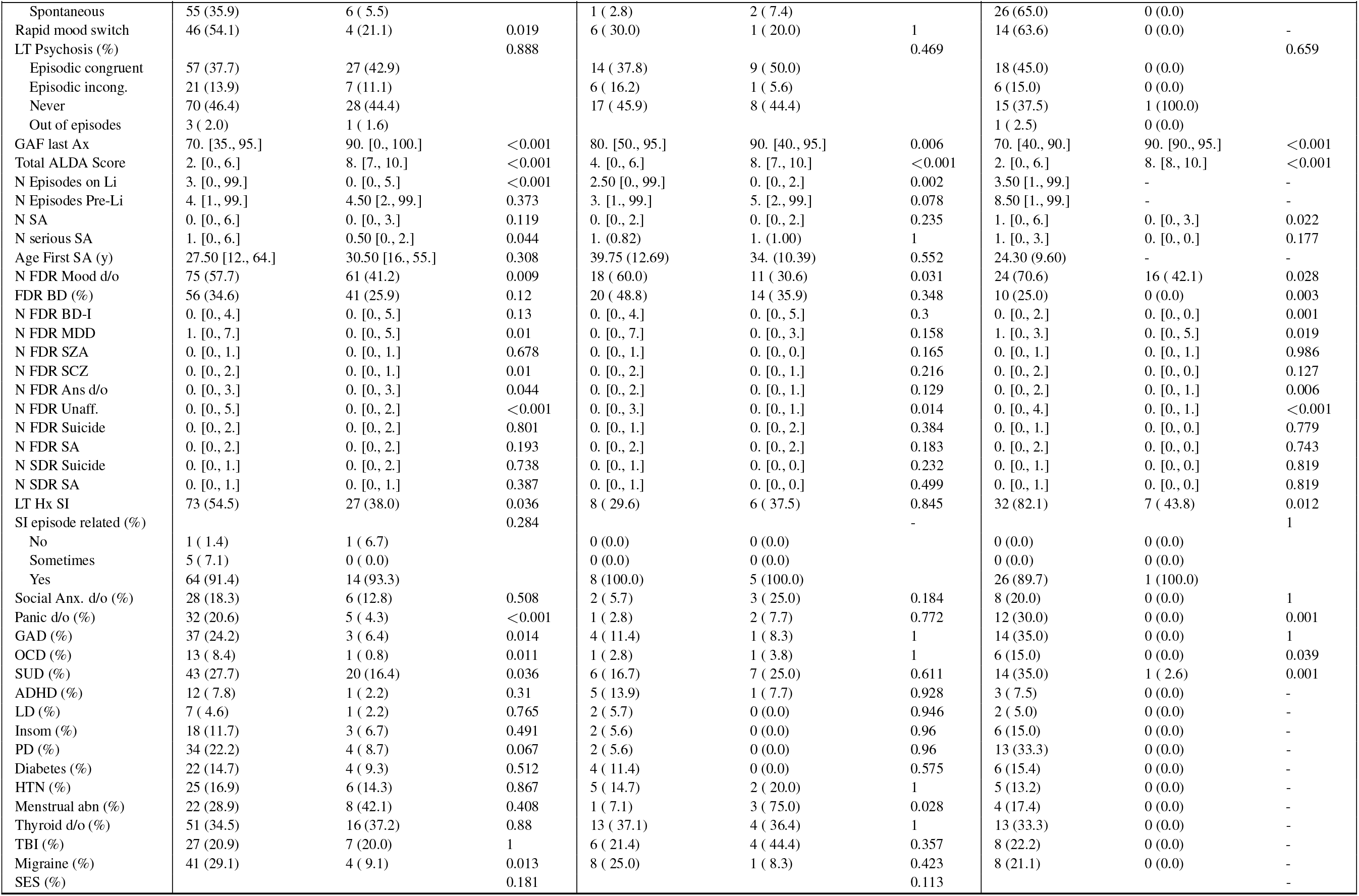
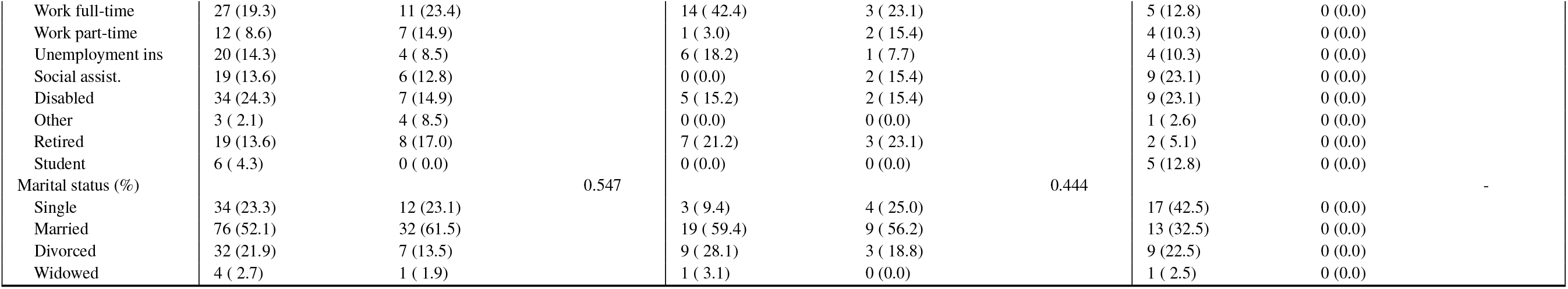
Demographic comparisons for subjects whose genomic data (from the Consortium for Lithium Genetics; ConLiGen) overlapped with our clinical dataset. Comparisons were done in between lithium responders (LR(+)) and non-responders (LR(-)) for the total group (“ALL), the best exemplars (“Best; exemplar score ≥ 75th percentile), and the poorest exemplars (“Poor; exemplar score ≤ 25th percentile).

**Table D.2.** Results of classifying lithium response based on the genomic data of all subjects (ALL; n=321), the poor exemplars (<25th percentile of exemplar score; n=81), and the best exemplars (>75th percentile of exemplar score; n=79). Each panel shows the results for a different classification performance metric. Classification was done using logistic regression with an L2 penalty (regularization weight set to C=1 a priori) with stratification done over each value of the resolution parameter q=1 and q=2. *Abbreviations*: accuracy (Acc), area under the receiver operating characteristic curve (AUC), sensitivity (Sens), specificity (Spec), Cohen’s kappa (Kappa), Matthews’ correlation coefficient (MCC), positive predictive value (PPV), negative predictive value (NPV). Results are presented as means and 95% confidence intervals.

| q | Group | Acc | AUC | Sens | Spec | PPV | NPV | Kappa | MCC |
| --- | --- | --- | --- | --- | --- | --- | --- | --- | --- |
| 1 | Best | 0.75 [0.66,0.87] | 0.88 [0.83,0.98] | 0.75 [0.50,0.94] | 0.88 [0.75,1.] | 0.90 [0.75,1.] | 0.71 [0.67,0.95] | 0.50 [0.31,0.74] | 0.58 [0.41,0.77] |
|  | Poor | 0.65 [0.53,0.75] | 0.66 [0.61,0.80] | 0.50 [0.31,0.75] | 0.75 [0.75,0.79] | 0.67 [0.53,0.75] | 0.67 [0.53,0.73] | 0.28 [0.06,0.50] | 0.29 [0.06,0.50] |
| 2 | Best | 0.75 [0.65,0.75] | 0.81 [0.66,0.86] | 0.75 [0.54,0.75] | 0.75 [0.56,0.94] | 0.75 [0.67,0.95] | 0.75 [0.67,0.79] | 0.50 [0.29,0.50] | 0.50 [0.39,0.58] |
|  | Poor | 0.50 [0.50,0.72] | 0.53 [0.45,0.72] | 0.50 [0.06,0.50] | 0.75 [0.50,1.] | 0.50 [0.12,0.90] | 0.50 [0.50,0.67] | 0. [0.00,0.44] | 0. [0.00,0.50] |
|  | ALL | 0.66 [0.60,0.70] | 0.70 [0.62,0.75] | 0.59 [0.48,0.62] | 0.70 [0.59,0.83] | 0.67 [0.59,0.78] | 0.65 [0.61,0.67] | 0.31 [0.20,0.39] | 0.32 [0.20,0.44] |

**Table D.3.**
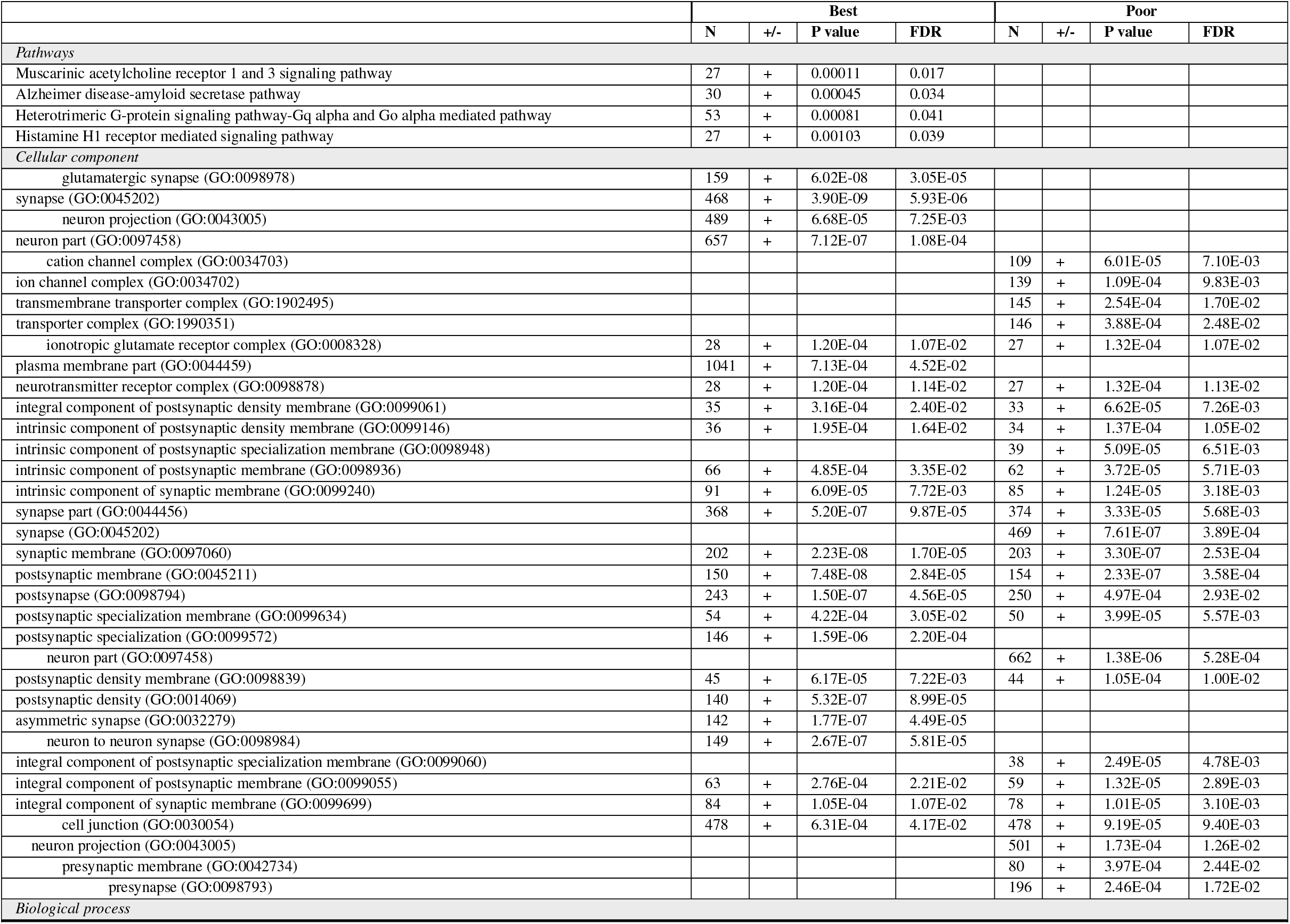
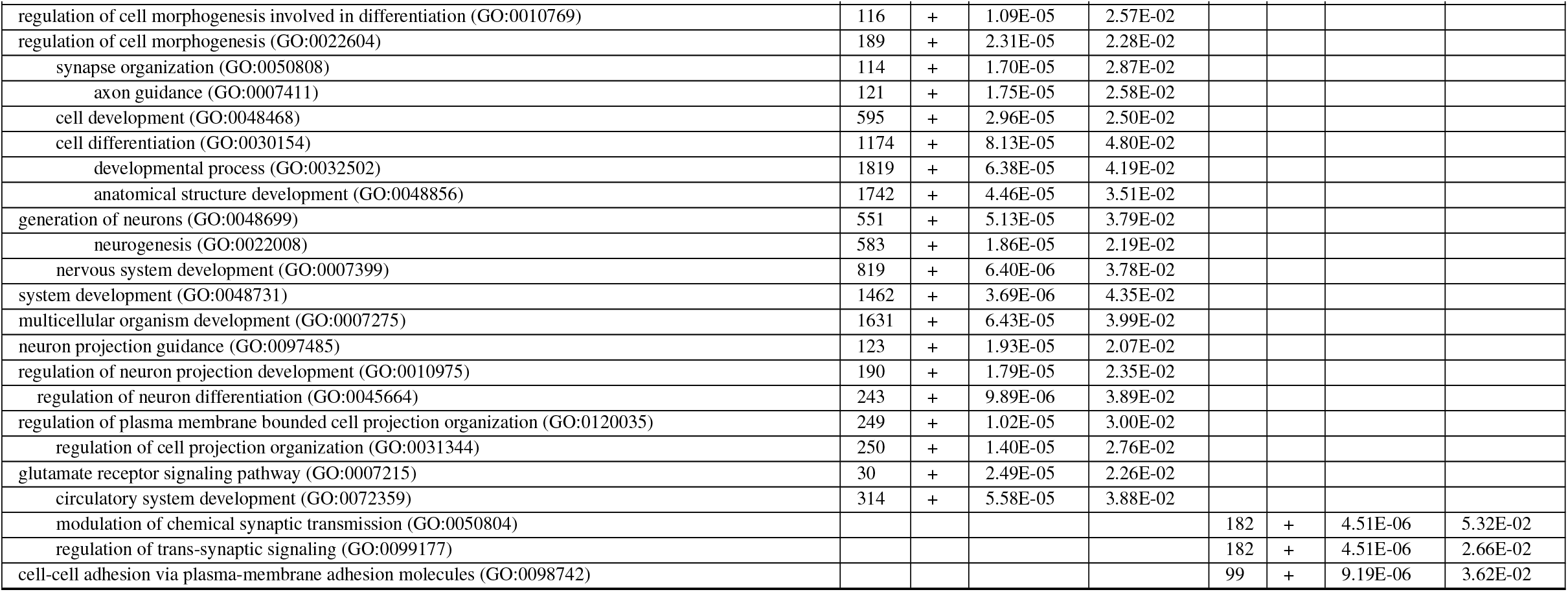
Results of gene enrichment analysis using the PANTHER gene ontology system. Analyses are presented for (A) pathways, (B) gene ontology cellular components, and (C) gene ontology biological processes. *Abbreviations*: false discovery rate (FDR).

**Table D.4.**
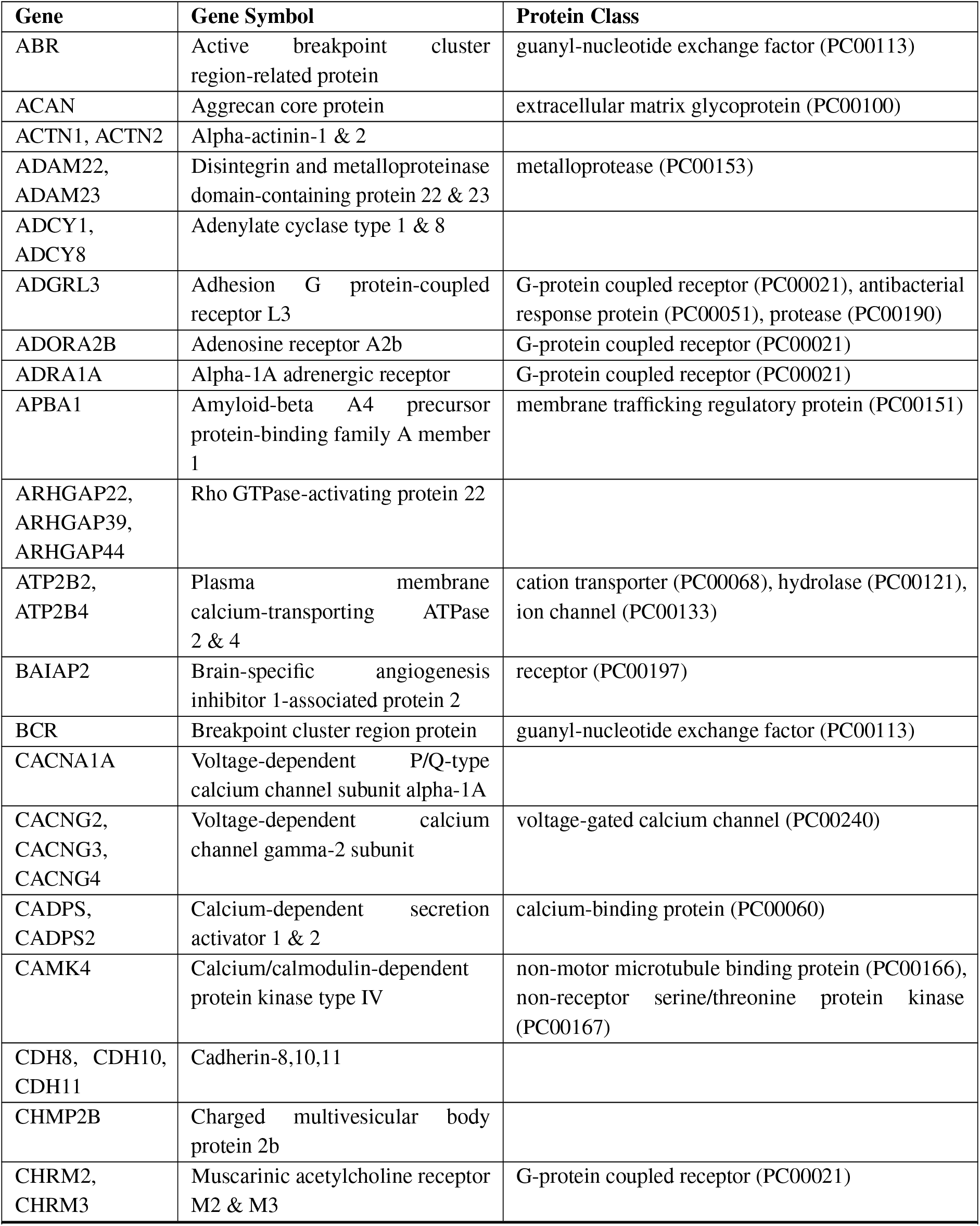
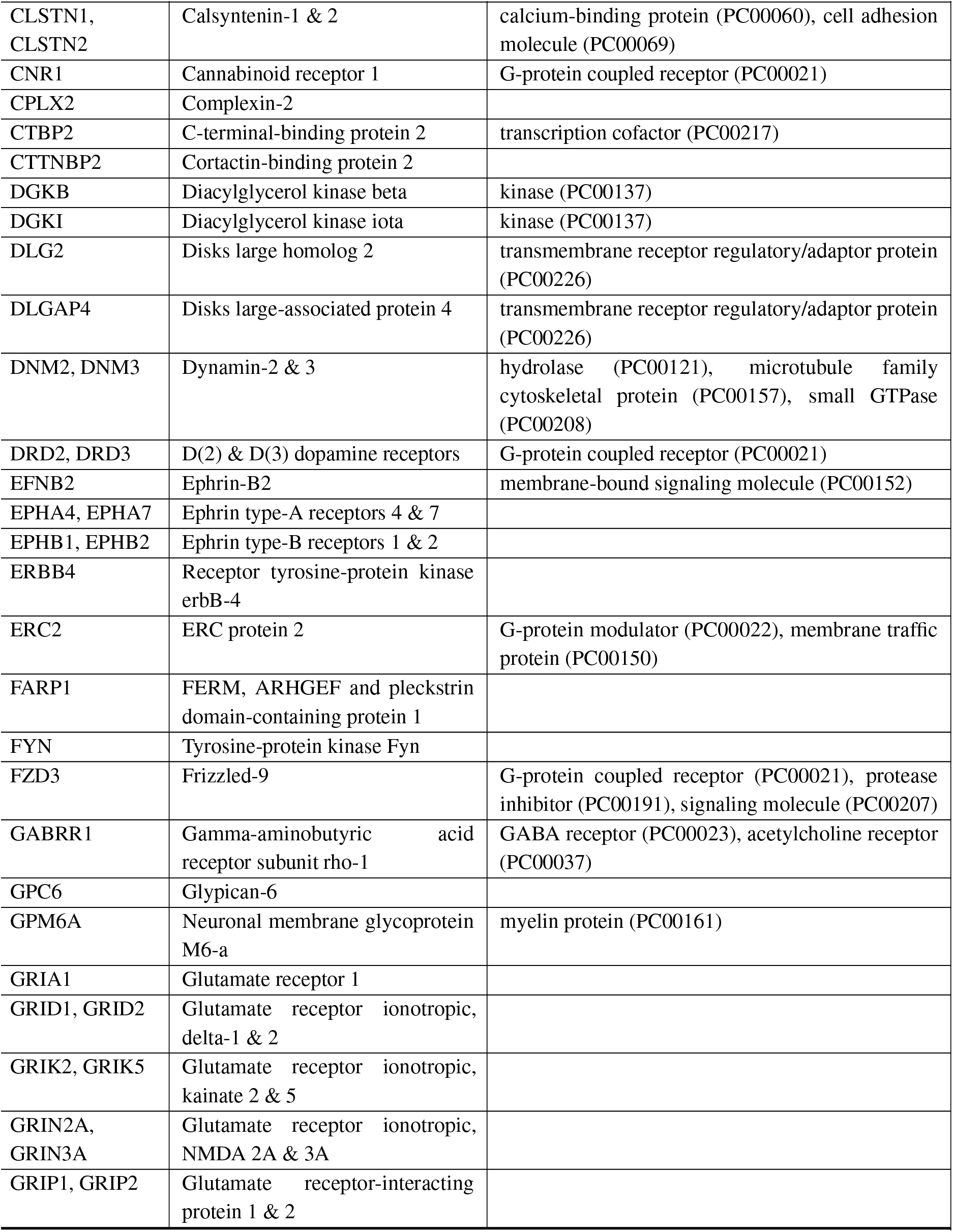
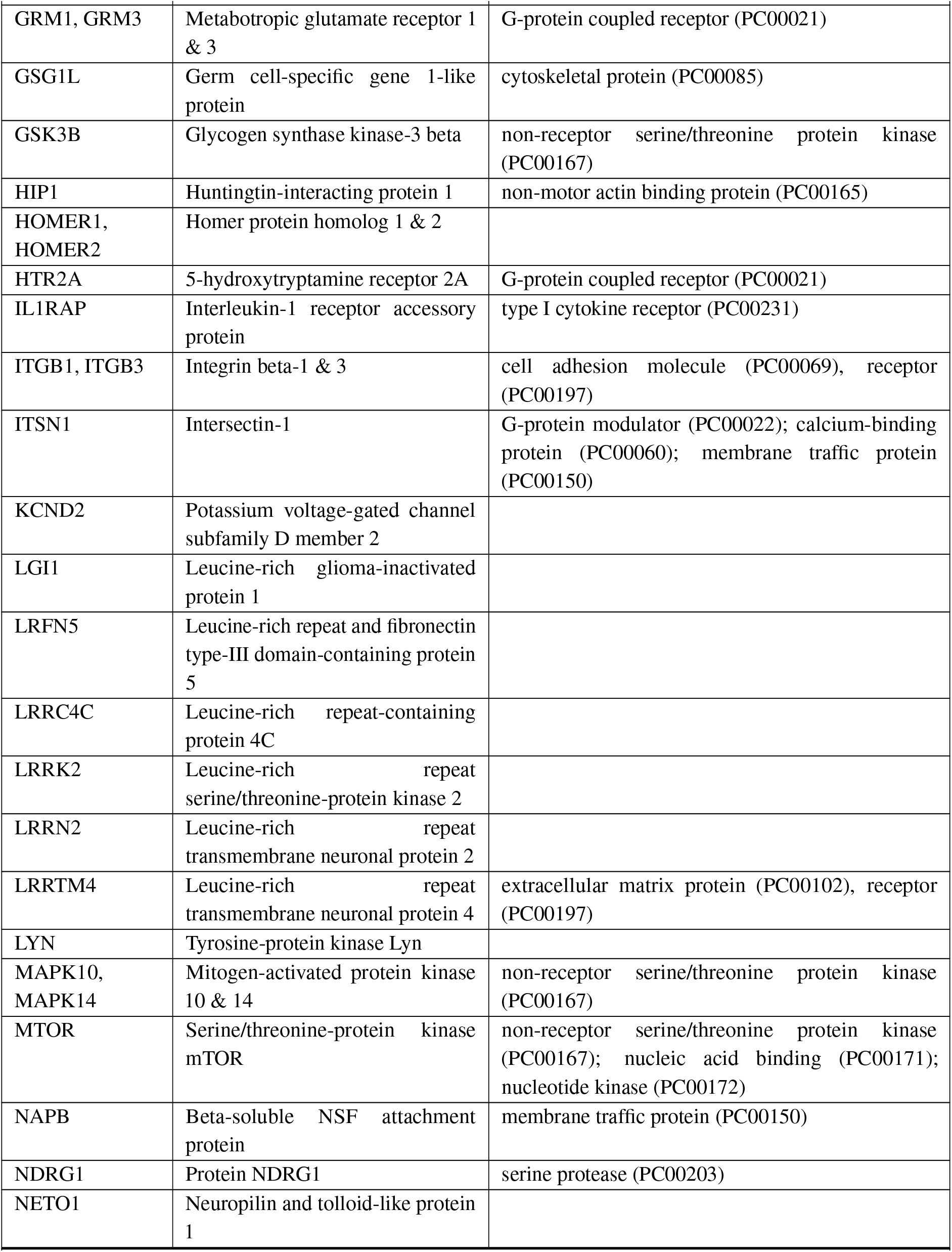
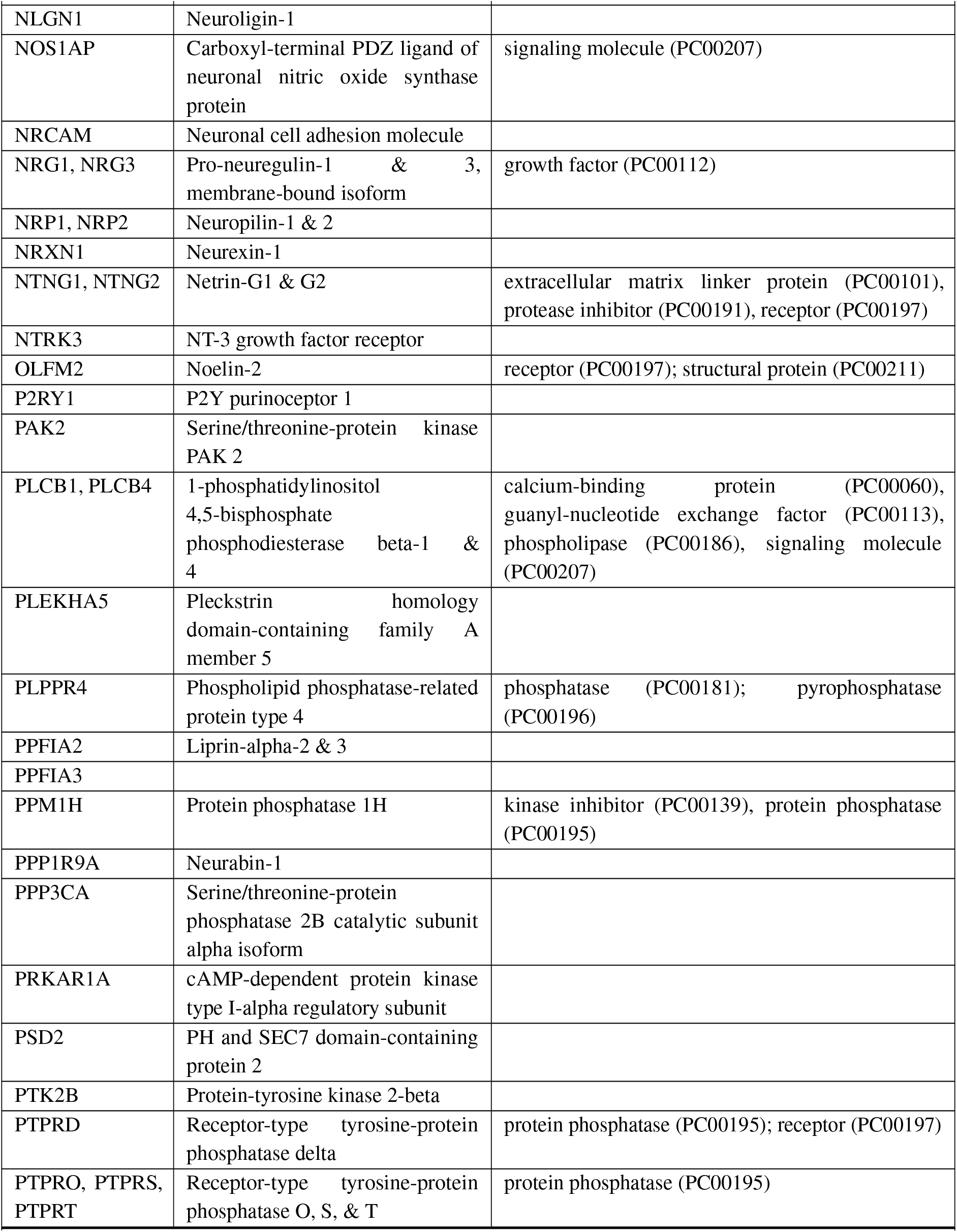
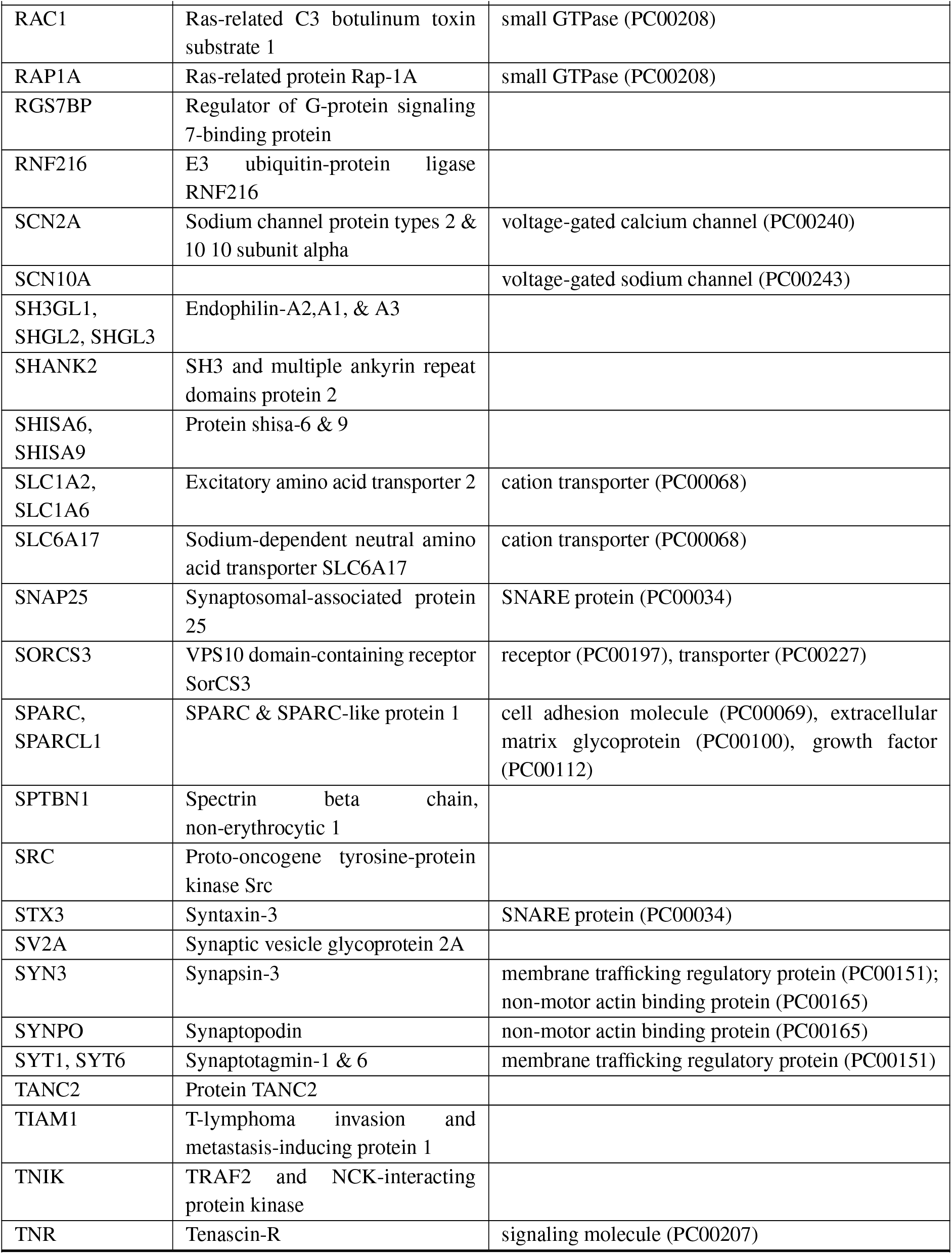
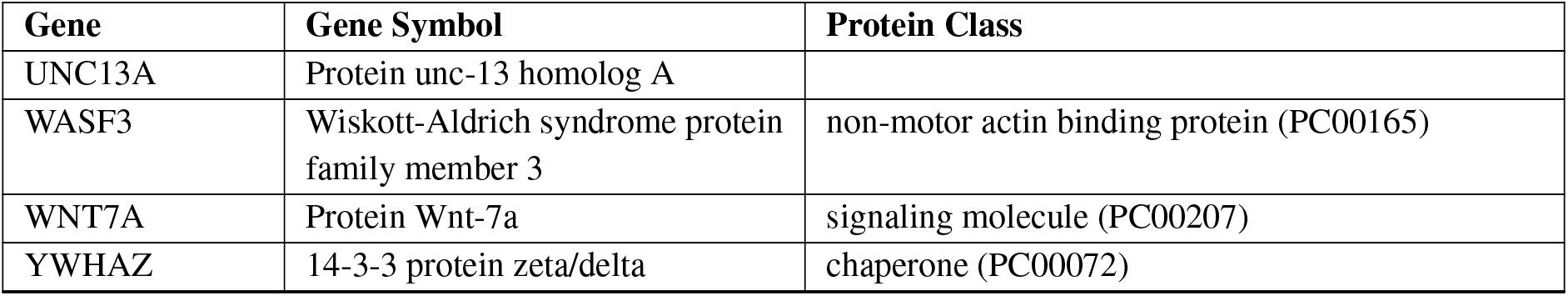
Genes enriched in the best exemplars group related to glutamatergic synapses (gene ontology “cellular component” category).

**Table D.5.**
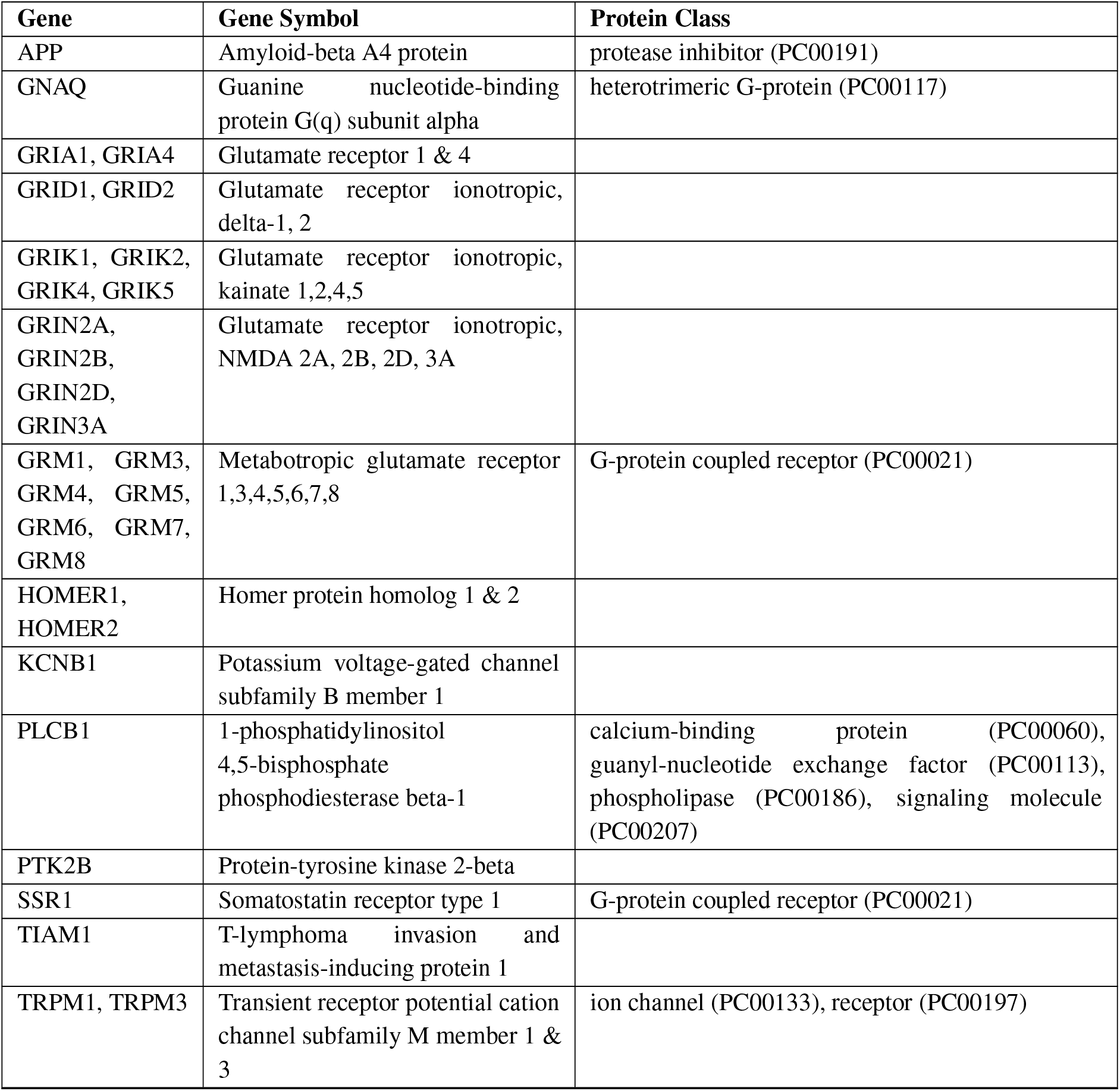
Genes enriched among the best exemplars in the gene ontology “biological process” category of the glutamate receptor signaling pathway.

| Gene | Gene Symbol | Protein Class |
| --- | --- | --- |
| APP | Amyloid-beta A4 protein | protease inhibitor (PC00191) |
| GNAQ | Guanine nucleotide-binding protein G(q) subunit alpha | heterotrimeric G-protein (PC00117) |
| GRIA1, GRIA4 | Glutamate receptor 1 & 4 |  |
| GRID1, GRID2 | Glutamate receptor ionotropic, delta-1, 2 |  |
| GRIK1, GRIK2, GRIK4, GRIK5 | Glutamate receptor ionotropic, kainate 1,2,4,5 |  |
| GRIN2A, GRIN2B, GRIN2D, GRIN3A | Glutamate receptor ionotropic, NMDA 2A, 2B, 2D, 3A |  |
| GRM1, GRM3, GRM4, GRM5, GRM6, GRM7, GRM8 | Metabotropic glutamate receptor 1,3,4,5,6,7,8 | G-protein coupled receptor (PC00021) |
| HOMER1, HOMER2 | Homer protein homolog 1 & 2 |  |
| KCNB1 | Potassium voltage-gated channel subfamily B member 1 |  |
| PLCB1 | 1-phosphatidylinositol 4,5-bisphosphate phosphodiesterase beta-1 | calcium-binding protein (PC00060), guanyl-nucleotide exchange factor (PC00113), phospholipase (PC00186), signaling molecule (PC00207) |
| PTK2B | Protein-tyrosine kinase 2-beta |  |
| SSR1 | Somatostatin receptor type 1 | G-protein coupled receptor (PC00021) |
| TIAM1 | T-lymphoma invasion and metastasis-inducing protein 1 |  |
| TRPM1, TRPM3 | Transient receptor potential cation channel subfamily M member 1 & 3 | ion channel (PC00133), receptor (PC00197) |

## Notes

### Competing Interest Statement

The authors have declared no competing interest.

